# Global Force-of-Infection Trends for Human *Taenia solium* Taeniasis/Cysticercosis

**DOI:** 10.1101/2022.02.11.22270710

**Authors:** Matthew A. Dixon, Peter Winskill, Wendy E. Harrison, Charles Whittaker, Veronika Schmidt, Astrid Carolina Flórez Sánchez, Zulma M. Cucunubá, Agnes U. Edia-Asuke, Martin Walker, Maria-Gloria Basáñez

## Abstract

Infection by *Taenia solium* poses a major burden across endemic countries. The World Health Organization (WHO) 2021–2030 Neglected Tropical Diseases roadmap has proposed that 30% of endemic countries achieve intensified *T. solium* control in hyperendemic areas by 2030. Understanding geographical variation in age-prevalence profiles and force-of-infection (FoI) estimates will inform intervention designs across settings.

Human taeniasis (HTT) and human cysticercosis (HCC) age-prevalence data from 16 studies in Latin America, Africa and Asia were extracted through a systematic review. Catalytic models, incorporating diagnostic performance uncertainty, were fitted to the data using Bayesian methods, to estimate rates of antibody (Ab)-seroconversion, infection acquisition and Ab-seroreversion or infection loss. HCC FoI and Ab-seroreversion rates were also estimated across 23 departments in Colombia from 28,100 individuals.

Across settings, there was extensive variation in all-ages seroprevalence. Evidence for Ab- seroreversion or infection loss was found in most settings for both HTT and HCC and for HCC Ab- seroreversion in Colombia. The average duration until humans became Ab-seropositive/infected decreased as all-age (sero)prevalence increased. There was no clear relationship between the average duration humans remain Ab-seropositive and all-age seroprevalence.

Marked geographical heterogeneity in *T. solium* transmission rates indicate the need for setting-specific intervention strategies to achieve the WHO goals.

## Introduction

The zoonotic cestode *Taenia solium* poses a substantial public health and economic challenge. Globally, *T. solium* is ranked as the foodborne parasitic infection contributing the highest number of disability-adjusted life-years (DALYs), an estimated 2.78 million DALYs in 2010 (Havelaar et al., 2015). Control and elimination of *T. solium* requires a One Health approach (Thomas et al., 2019; Braae et al., 2019) due to its complex multi-host life-cycle, involving infection with larval-stage cysticerci in the intermediate pig host (porcine cysticercosis, or PCC), infective stages (eggs) in the environment, and taeniasis (infection with the adult worm) in the definitive human host (human taeniasis, or HTT). When humans ingest *T. solium* eggs, establishment of cysticerci (human cysticercosis, or HCC) in the central nervous system results in neurocysticercosis (NCC). Neurocysticercosis is responsible for the major health burden associated with *T. solium*, causing approximately one third of epilepsy/seizure disorders in endemic settings (Gripper and Welburn, 2017).

Current interventions target PCC through mass treatment of pigs with oxfendazole and their vaccination with TSOL18, and HTT through mass treatment of humans with praziquantel or niclosamide. These control strategies have proven efficacious in field studies (de Coster et al., 2018; Dixon et al., 2021), but to date no large-scale interventions have been rolled out as part of national Neglected Tropical Disease (NTD) programmes, and therefore their impact alone or in combination in different epidemiological settings remains poorly understood and/or modelled (CystiTeam, 2019). The milestones proposed in the new World Health Organization (WHO) NTD 2021–2030 roadmap (World Health Organization, 2020) focus on achieving intensified control in hyperendemic areas of 17 (27%) endemic countries. However, endemicity levels for *T. solium* have not been defined in terms of infection indicators and there exist limited data on the geographical distribution of *T. solium* prevalence at national scales. For other NTDs, pre-intervention levels of endemicity (according to infection prevalence, intensity, incidence or morbi-mortality) determine the magnitude, duration and likely success of control efforts (NTD Modelling Consortium Onchocerciasis Group, 2019). Therefore, successful *T. solium* intervention strategies will require tailoring to local epidemiological and socio-economic conditions that determine the intensity of transmission and will likely be informed by age- and/or sex-dependent contact rates and mixing patterns in endemic settings (Dixon et al., 2021; CystiTeam, 2019, Welburn et al., 2015).

Global patterns in transmission rates have recently been explored for PCC by assessing the force-of-infection (FoI; the per-capita rate of infection acquisition) according to proposed endemicity levels (overall (sero)prevalence) and geography (Dixon et al., 2020). Estimates of antibody (Ab) and antigen (Ag) seroconversion (and seroreversion rates) for HCC have also been estimated from a small number of longitudinal surveys in Burkina Faso (Dermauw et al., 2018), Peru (Garcia et al., 2001)], Ecuador (Coral-Almeida et al., 2014) and Zambia (Mwape et al., 2013), demonstrating substantial geographic variation.

Age-prevalence profiles are not only useful for estimating FoI and (sero)reversion rates, but also to provide insight into processes driving epidemiological dynamics, such as immunological responses or heterogeneity in exposure which may explain abrupt changes in HCC seropositivity in older individuals or distinct prevalence peaks in younger ages for HTT prevalence, as observed in the Democratic Republic of the Congo (DRC) (Kanobana et al., 2011; Madinga et al., 2017). Specific age-prevalence profiles may also provide insight into whether age-targeted interventions may be appropriate in certain settings. This study, therefore, and for the first time systematically collates and reviews all HTT and HCC age-(sero)prevalence data available from the literature and from collaborators, and uses simple and reversible catalytic models to estimate the FoI and Ab- seroreversion/infection loss rates from cross-sectional surveys. The results improve our understanding of global variation in the epidemiology of *T. solium* and provide estimates of transmission rates, crucial for guiding the design of interventions. In addition, a country profile of FoI estimates is presented at the sub-national level for Colombia to exemplify how detailed FoI analyses can help understand local epidemiological patterns.

## Results

### Systematic review and study selection

After title, abstract and full-text eligibility screening of 236 studies initially identified (01/11/2014 to 02/10/2019), and 11 studies included in the Coral-Almeida et al. (2015) literature review, a total of 16 studies were included in the analysis (PRISMA flowchart; Supplementary File 1: Supplementary Figure S1), originating from South America (*n* = 4), Africa (*n* = 8) and Asia (*n* = 4) (Supplementary File 1: Supplementary Figure S2) and split by *n* = 4 HTT and *n* = 15 HCC surveys (full details in Supplementary File 1: Supplementary Table S1). Total sample sizes were 6,653 individuals (range 576–4,599; with individual age range of <1 to 96 years) across HTT surveys, which included three copro-Ag-based surveys and one Ab-based survey; 34,124 (125–29,360; cross-study age range of <1 to 95 years) across HCC-Ab surveys, and 12,934 (905–4,993; cross-study age range of <1 to 96 years) across HCC-Ag surveys. Observed (sero)prevalence ranged from 4.5% to 23.4% (95% confidence interval (CI) range: 3.0%–24.6%) for HTT surveys, 0.5%–38.7% (0.1%–41.6%) across HCC-Ab surveys, and 0.7%–21.7% (0.5%–24.5%) across HCC-Ag surveys.

Catalytic models (Figure 1; full details in Material and methods) were fitted to the thus extracted (sero)prevalence data using a Bayesian framework that integrates uncertainty in diagnostic sensitivity and specificity from prior (published) information (Supplementary File 1: Supplementary Table S1). The FoI parameters of acquisition (*λ*) and reversion/loss (*ρ*) (for antibody datasets: *λ*_*sero*_ and *ρ*_*sero*_; for antigen datasets: *λ*_*inf*_ and *ρ*_*inf*_) were estimated for each dataset. The deviance information criterion (DIC) (Spiegelhalter et al., 2002) was used to compare individually- and jointly-fitted catalytic models.

**Figure 1.**
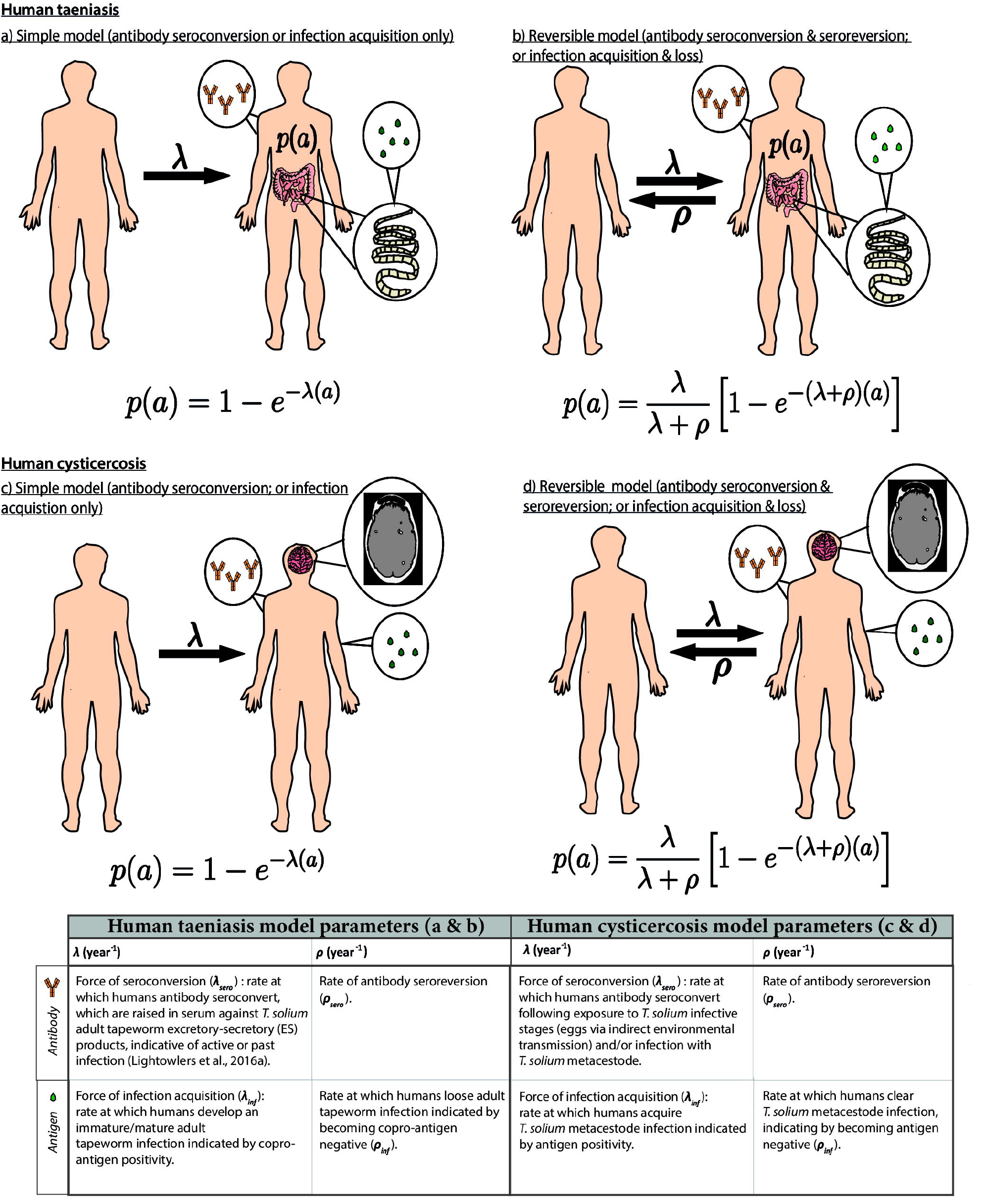
Simple and reversible catalytic model structure and equations fitted to data on the age (a)-specific (sero)prevalence (*p*(*a*)) data, where *λ* is the force-of-infection (rate of antibody (Ab)- seroconversion or infection acquisition) and *ρ* the rate of Ab-seroreversion or infection loss. The general mathematical form of the catalytic models (equations 1a to 1d) fitted to the human taeniasis (HTT)/human cysticercosis (HCC) Ab, HCC Ag and HTT copro-Ag prevalence datasets to estimate the prevalence (*p*) at human age (*a*). The saturating (sero)prevalence is given by *λ*/(*λ* + *ρ*) which for the simple model is 100%, if the humans lived sufficiently long. The accompanying tables provide information on the definitions of the catalytic model parameters depending on the diagnostic method. Presence of adult tapeworm excretory-secretory products indicative of active or past HTT infection, as outlined by Lightowlers et al. 2016a.

We defined hyperendemic transmission settings as those with all-age *observed* HTT (sero)prevalence of ≥3% and all-age *observed* HCC (sero)prevalence of ≥6%. Studies with (sero)prevalence values below these were defined as endemic transmission settings. These putative endemicity definitions were defined following a literature review of studies referring to “hyper” or “highly” endemic settings (Supplementary File 1: Supplementary Table S2).

### Global human taeniasis (HTT) copro-antigen and antibody seroprevalence

Table 1 compares models fitted either including (reversible model) or excluding (simple model) HTT infection loss (when fitted to copro-Ag ELISA using the Allan et al. (1990) protocol datasets, except in Gomes et al. (2002) where a protocol is not specified) or Ab-seroreversion. For the copro-Ag ELISA datasets (to which models were jointly fitted to yield a single sensitivity and specificity posterior), DIC scores were similar (within one unit) between models with and without infection loss (Table 1 and Figure 2a), indicating limited information to differentiate between model fits.

**Table 1.**
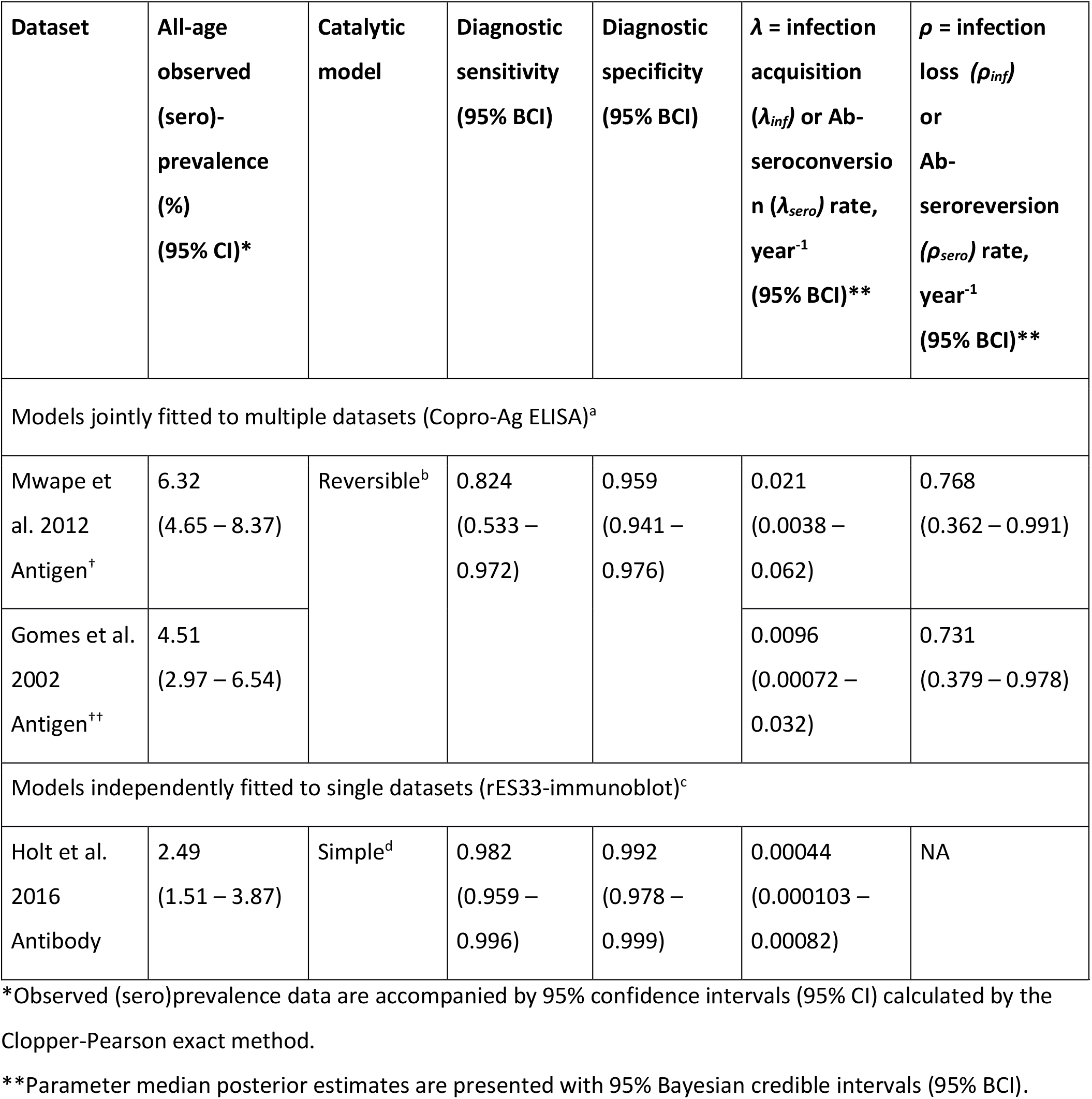
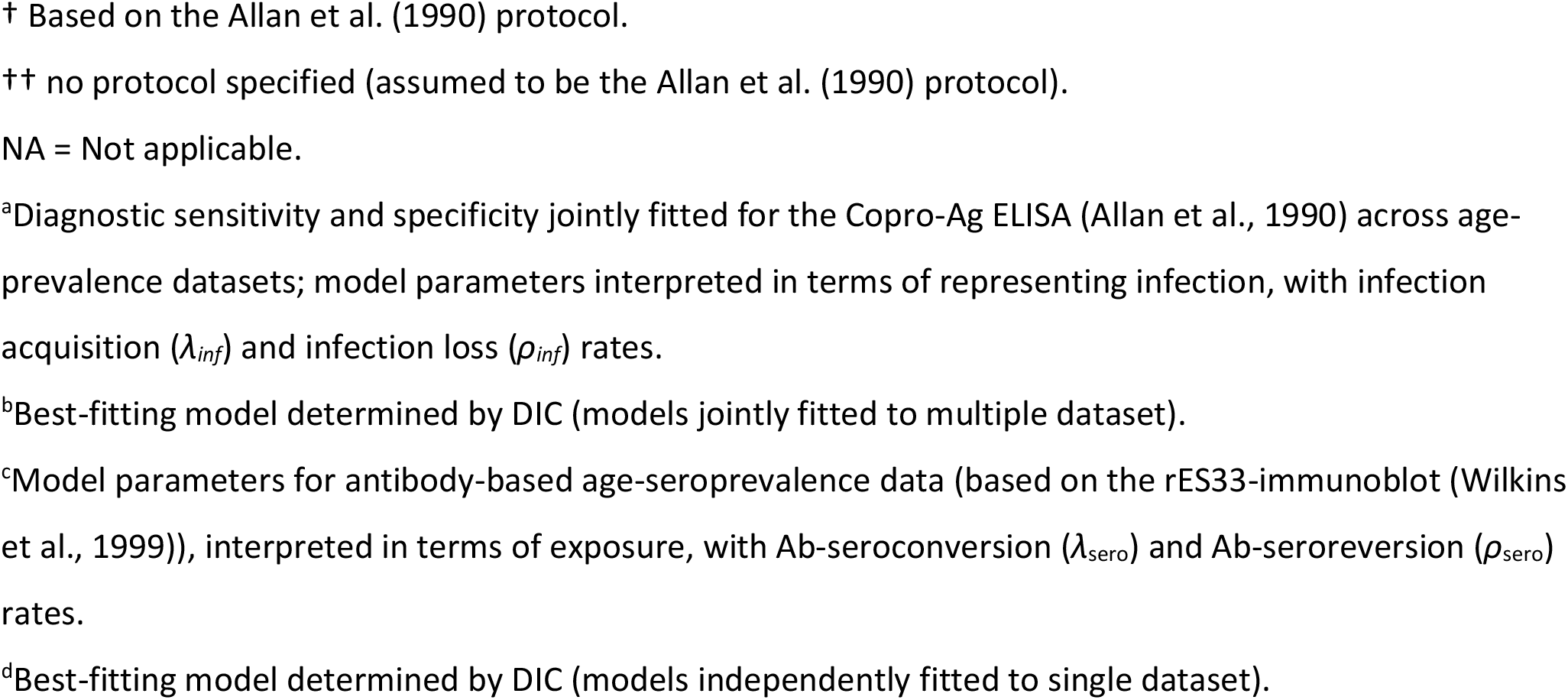
(Sero)prevalence and parameter posterior estimates for the best-fit catalytic models fitted to human taeniasis (antibody and antigen) age-(sero)prevalence datasets (ordered by decreasing all-age (sero)prevalence).

| Dataset | All-age<br>observed<br>(sero)-<br>prevalence<br>(%)<br>(95% CI)* | Catalytic<br>model | Diagnostic<br>sensitivity<br>(95% BCI) | Diagnostic<br>specificity<br>(95% BCI) | $\lambda$ = infection<br>acquisition<br>($\lambda_{inf}$) or Ab-<br>seroconver-<br>sion ($\lambda_{sero}$) rate,<br>year<sup>-1</sup><br>(95% BCI)** | $\rho$ = infection<br>loss ($\rho_{inf}$)<br>or<br>Ab-seroreversion<br>($\rho_{sero}$) rate,<br>year<sup>-1</sup><br>(95% BCI)** |
| --- | --- | --- | --- | --- | --- | --- |
| Models jointly fitted to multiple datasets (Copro-Ag ELISA)<sup>a</sup> |  |  |  |  |  |  |
| Mwape et<br>al. 2012<br>Antigen<sup>†</sup> | 6.32<br>(4.65 – 8.37) | Reversible<sup>b</sup> | 0.824<br>(0.533 –<br>0.972) | 0.959<br>(0.941 –<br>0.976) | 0.021<br>(0.0038 –<br>0.062) | 0.768<br>(0.362 – 0.991) |
| Gomes et al.<br>2002<br>Antigen<sup>††</sup> | 4.51<br>(2.97 – 6.54) |  |  |  | 0.0096<br>(0.00072 –<br>0.032) | 0.731<br>(0.379 – 0.978) |
| Models independently fitted to single datasets (rES33-immunoblot)<sup>c</sup> |  |  |  |  |  |  |
| Holt et al.<br>2016<br>Antibody | 2.49<br>(1.51 – 3.87) | Simple<sup>d</sup> | 0.982<br>(0.959 –<br>0.996) | 0.992<br>(0.978 –<br>0.999) | 0.00044<br>(0.000103 –<br>0.00082) | NA |

\*Observed (sero)prevalence data are accompanied by 95% confidence intervals (95% CI) calculated by the Clopper-Pearson exact method.
\*\*Parameter median posterior estimates are presented with 95% Bayesian credible intervals (95% BCI).
† Based on the Allan et al. (1990) protocol.
†† no protocol specified (assumed to be the Allan et al. (1990) protocol).
NA = Not applicable.
<sup>a</sup>Diagnostic sensitivity and specificity jointly fitted for the Copro-Ag ELISA (Allan et al., 1990) across age-prevalence datasets; model parameters interpreted in terms of representing infection, with infection acquisition ( $\lambda_{inf}$ ) and infection loss ( $\rho_{inf}$ ) rates.
<sup>b</sup>Best-fitting model determined by DIC (models jointly fitted to multiple dataset).
<sup>c</sup>Model parameters for antibody-based age-seroprevalence data (based on the rES33-immunoblot (Wilkins et al., 1999)), interpreted in terms of exposure, with Ab-seroconversion ( $\lambda_{sero}$ ) and Ab-seroreversion ( $\rho_{sero}$ ) rates.
<sup>d</sup>Best-fitting model determined by DIC (models independently fitted to single dataset).

**Figure 2.**
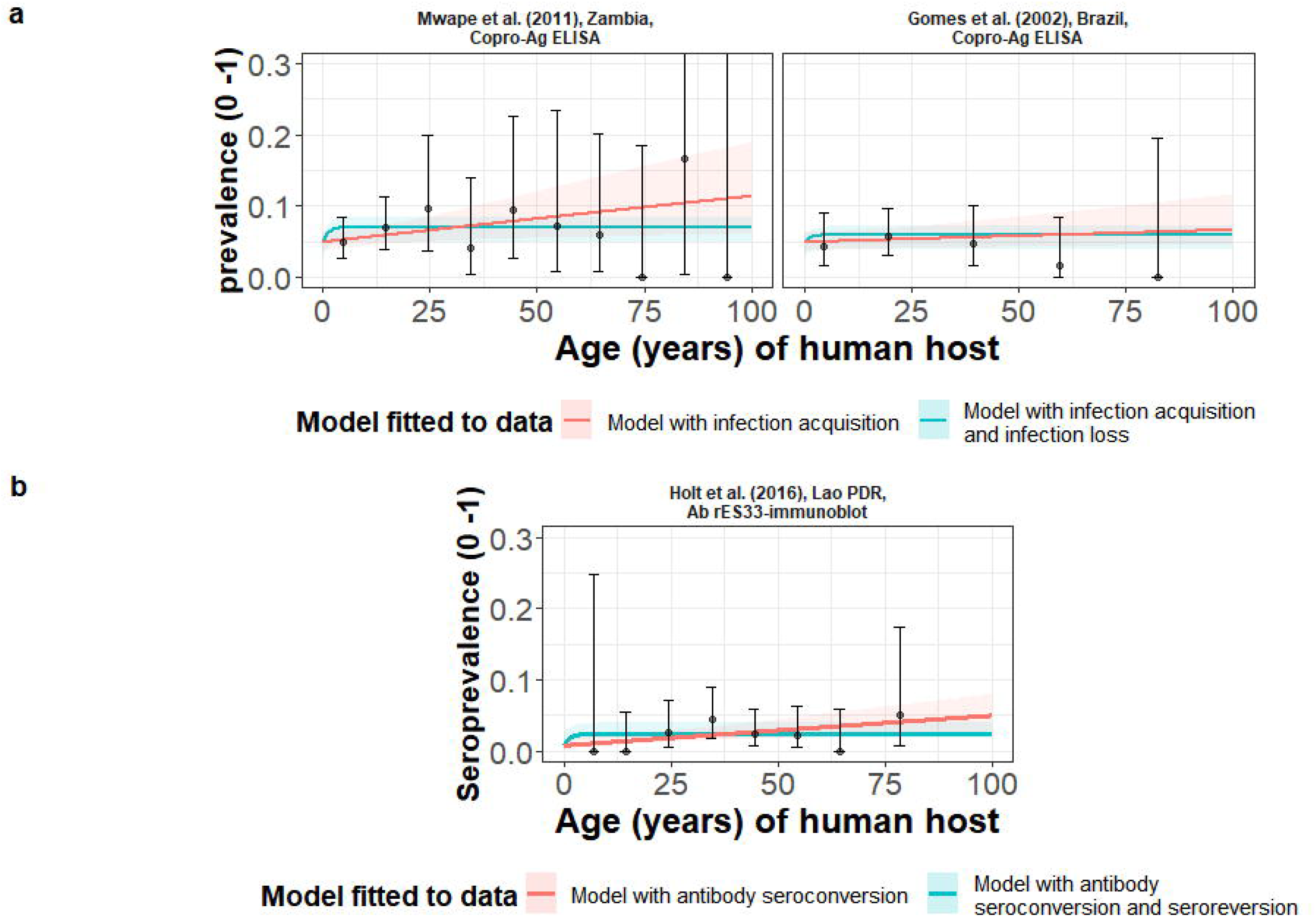
The relationship between human taeniasis copro-antigen (Copro-Ag) and antibody (sero)prevalence and human age (in years) for each dataset. Human taeniasis (HTT) infection acquisition (simple) or acquisition with infection loss (reversible) catalytic models jointly fitted to multiple datasets (where single diagnostic sensitivity and specificity values were estimated; dataset-specific *λ* and *ρ* estimates were obtained) in **a**); HTT antibody (Ab)-seroconversion (simple) or Ab- seroconversion with Ab-seroreversion to a single dataset in **b**). 95% confidence intervals associated with observed (sero)prevalence point estimates are also presented. Bayesian Markov chain Monte Carlo methods were used to fit the models to data, with the parameter posterior distributions used to construct predicted (all age) (sero)prevalence curves and associated 95% Bayesian credible intervals (BCIs). Best-fitting model selected by deviance information criterion (DIC); both models presented if difference between DIC < 2 (both models have similar support based on the data); a difference > 10 units indicates that the models are significantly different and therefore only superior fitting model (lowest DIC) is presented). The non-zero predicted (sero)prevalence at age 0 is due to less than 100% specificity for all tests. The 95% confidence intervals (95% CI) for age-(sero)prevalence data-points are calculated by the Clopper-Pearson exact method.

The copro-Ag ELISA datasets (Gomes et al., 2002; Mwape et al., 2012) were found in hyperendemic settings (all-age HTT (sero)prevalence ≥3%) in Zambia (6.3%) (Mwape et al., 2012) and Brazil (4.5%) (Gomes et al., 2002). For the models independently fitted to the single dataset in Lao People’s Democratic Republic (Lao PDR) (Holt et al., 2016) using the rES33-immunoblot (Wilkin et al., 1999) and found in an endemic setting (all-age HTT antibody seroprevalence of 2.5%), there was also limited information to differentiate between model fits, with DIC scores similar (within one unit) between the model with and without Ab-seroconversion (Table 1 and Figure 2b). The FoI (*λ*_*sero*_) for the best-fit model to the rES33-immunoblot antibody dataset in Lao PDR (Holt et al., 2016), suggested a very low HTT Ab-seroconversion rate of 0.00046 year^-1^ (all model fits and DIC scores in Supplementary File 1: Supplementary File Table S3). The Madinga et al. (2017) dataset in the DRC was omitted from the models jointly fitted across copro-Ag ELISA datasets, due to difficulty fitting catalytic models to such a distinct age-prevalence profile with a marked peak in early ages (see Discussion).

### Global human cysticercosis (HCC) antibody seroprevalence

HCC Ab-seroconversion with Ab-seroreversion (reversible model) provided an improved joint fit to the multiple datasets based on the antibody lentil lectin-purified glycoprotein enzyme-linked immunoelectrotransfer blot (LLGP-EITB) assay (Tsang et al., 1989) (Table 2 and Figure 3a). These fits were found in the proposed hyperendemic settings (≥6% HCC seroprevalence; with all-age seroprevalence from 12.7% to 24.7%) in Peru (Lescano et al., 2009; Moro et al., 2003), India (Jayaraman et al., 2011), and Bali (Theis et al., 1994), and an endemic setting in Brazil (1.6%) (Gomes et al., 2002). For the models jointly fitted to multiple datasets based on the IgG Ab-ELISA (*DiagAutom*, 2016) hyperendemic setting: all-age seroprevalence from 9.6% to 14.5%) in Nigeria (Edia-Asuke et al., 2015; Weka et al., 2013), and the models independently fitted to the single dataset from Lao PDR (endemic setting: 3.0% all-age seroprevalence) (Holt et al., 2016) based on the rT24H-immunoblot (Hancock et al., 2006), the (simple) model without Ab-seroreversion provided an improved fit (Table 2 and Figure 3b and 3c). Supplementary File 1: Supplementary File Table S4 presents all model fits and DIC scores.

**Table 2.**
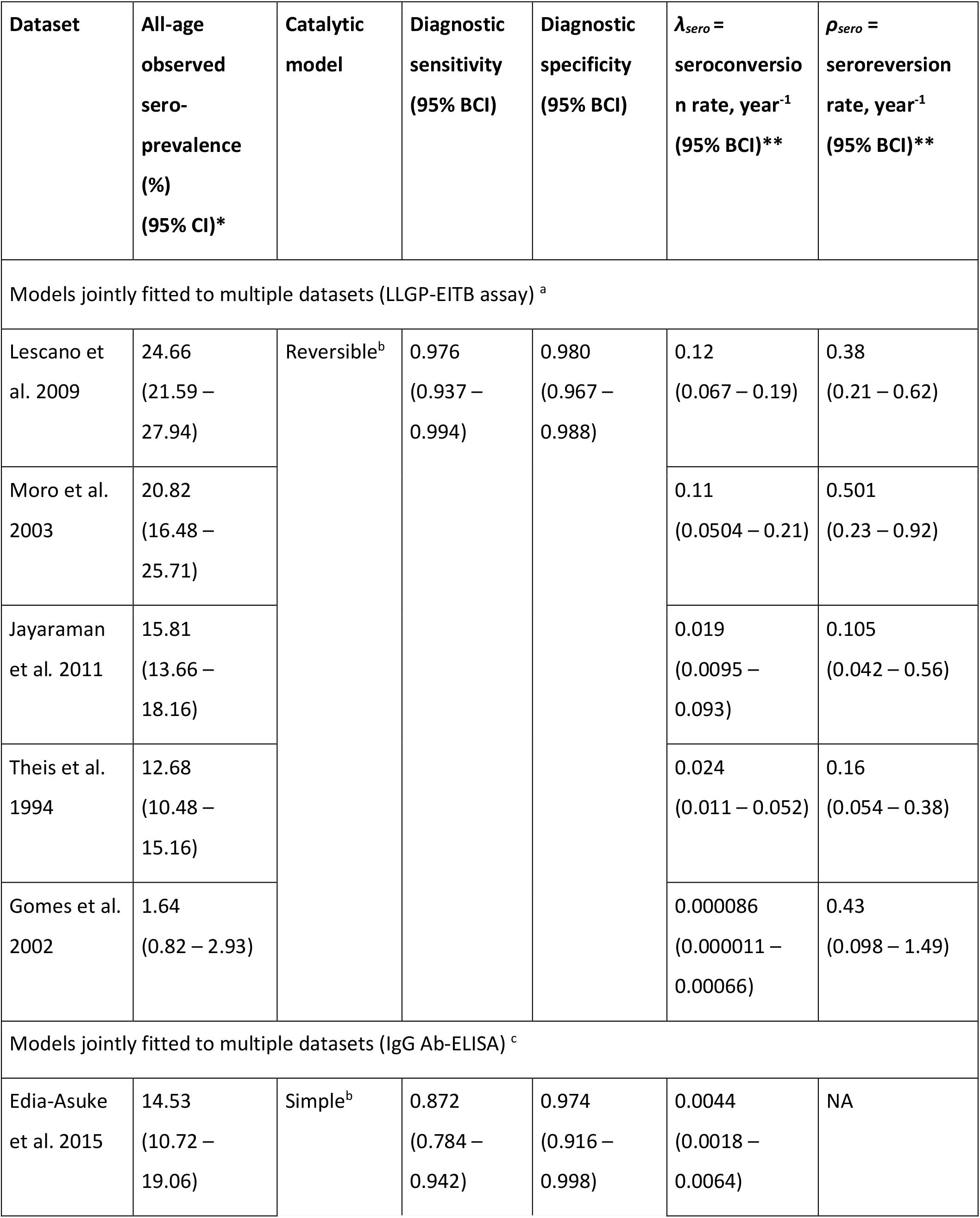
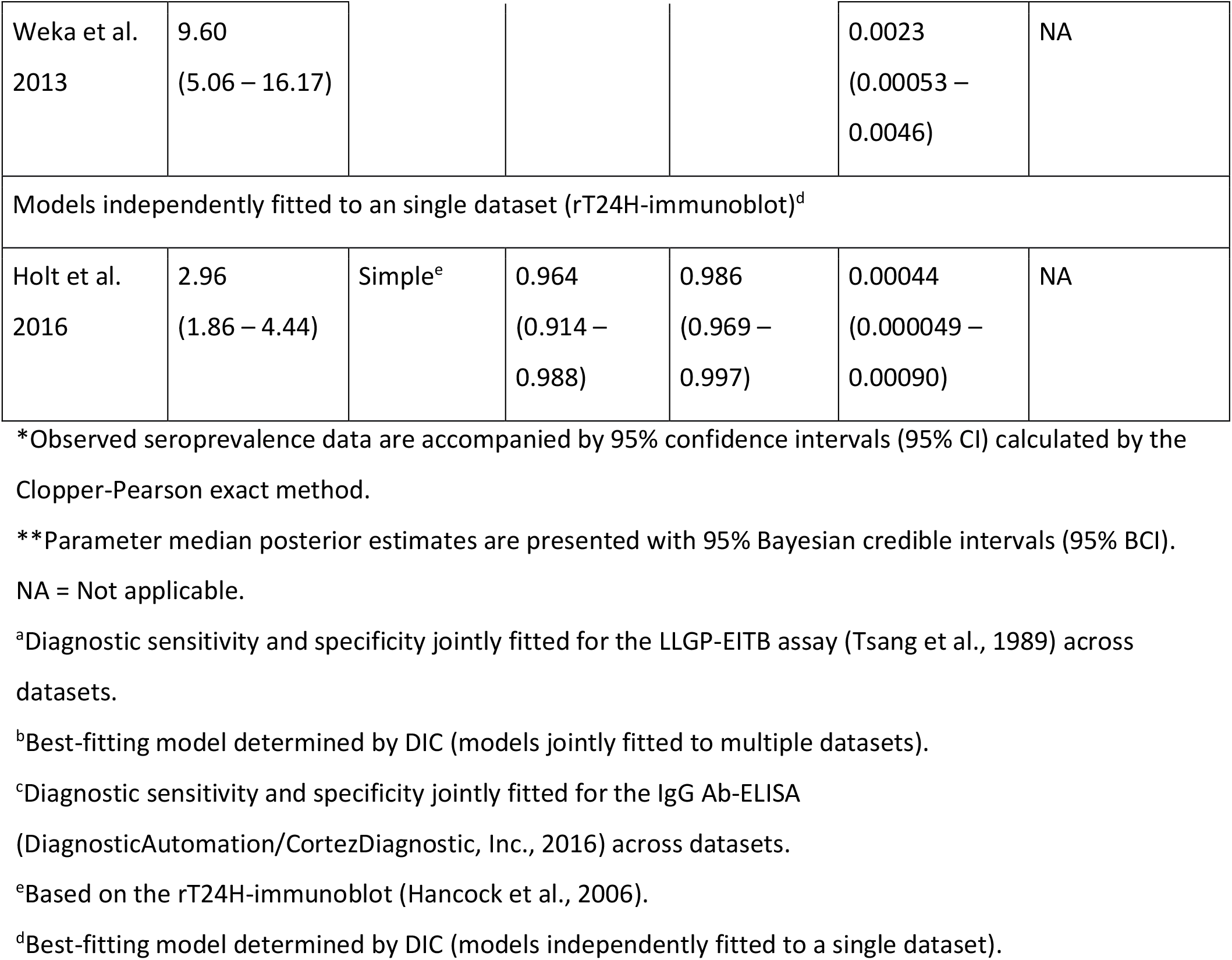
Antibody seroprevalence and parameter estimates for the best-fit catalytic models fitted to each observed human cysticercosis (antibody) age-seroprevalence dataset (ordered by decreasing all-age seroprevalence).

| Dataset | All-age<br>observed<br>sero-<br>prevalence<br>(%)<br>(95% CI)* | Catalytic<br>model | Diagnostic<br>sensitivity<br>(95% BCI) | Diagnostic<br>specificity<br>(95% BCI) | $\lambda_{sero}$ =<br>seroconversio<br>n rate, year<sup>-1</sup><br>(95% BCI)** | $\rho_{sero}$ =<br>seroreversion<br>rate, year<sup>-1</sup><br>(95% BCI)** |
| --- | --- | --- | --- | --- | --- | --- |
| Models jointly fitted to multiple datasets (LLGP-EITB assay) <sup>a</sup> |  |  |  |  |  |  |
| Lescano et al. 2009 | 24.66<br>(21.59 – 27.94) | Reversible<sup>b</sup> | 0.976<br>(0.937 – 0.994) | 0.980<br>(0.967 – 0.988) | 0.12<br>(0.067 – 0.19) | 0.38<br>(0.21 – 0.62) |
| Moro et al. 2003 | 20.82<br>(16.48 – 25.71) |  |  |  | 0.11<br>(0.0504 – 0.21) | 0.501<br>(0.23 – 0.92) |
| Jayaraman et al. 2011 | 15.81<br>(13.66 – 18.16) |  |  |  | 0.019<br>(0.0095 – 0.093) | 0.105<br>(0.042 – 0.56) |
| Theis et al. 1994 | 12.68<br>(10.48 – 15.16) |  |  |  | 0.024<br>(0.011 – 0.052) | 0.16<br>(0.054 – 0.38) |
| Gomes et al. 2002 | 1.64<br>(0.82 – 2.93) |  |  |  | 0.000086<br>(0.000011 – 0.00066) | 0.43<br>(0.098 – 1.49) |
| Models jointly fitted to multiple datasets (IgG Ab-ELISA) <sup>c</sup> |  |  |  |  |  |  |
| Edia-Asuke et al. 2015 | 14.53<br>(10.72 – 19.06) | Simple<sup>b</sup> | 0.872<br>(0.784 – 0.942) | 0.974<br>(0.916 – 0.998) | 0.0044<br>(0.0018 – 0.0064) | NA |
| Weka et al.<br>2013 | 9.60<br>(5.06 – 16.17) |  |  |  | 0.0023<br>(0.00053 –<br>0.0046) | NA |
| Models independently fitted to an single dataset (rT24H-immunoblot)<sup>d</sup> |  |  |  |  |  |  |
| Holt et al.<br>2016 | 2.96<br>(1.86 – 4.44) | Simple<sup>e</sup> | 0.964<br>(0.914 –<br>0.988) | 0.986<br>(0.969 –<br>0.997) | 0.00044<br>(0.000049 –<br>0.00090) | NA |

\*Observed seroprevalence data are accompanied by 95% confidence intervals (95% CI) calculated by the Clopper-Pearson exact method.
\*\*Parameter median posterior estimates are presented with 95% Bayesian credible intervals (95% BCI).
NA = Not applicable.
<sup>a</sup>Diagnostic sensitivity and specificity jointly fitted for the LLGP-EITB assay (Tsang et al., 1989) across datasets.
<sup>b</sup>Best-fitting model determined by DIC (models jointly fitted to multiple datasets).
<sup>c</sup>Diagnostic sensitivity and specificity jointly fitted for the IgG Ab-ELISA (DiagnosticAutomation/CortezDiagnostic, Inc., 2016) across datasets.
<sup>e</sup>Based on the rT24H-immunoblot (Hancock et al., 2006).
<sup>d</sup>Best-fitting model determined by DIC (models independently fitted to a single dataset).

**Figure 3.**
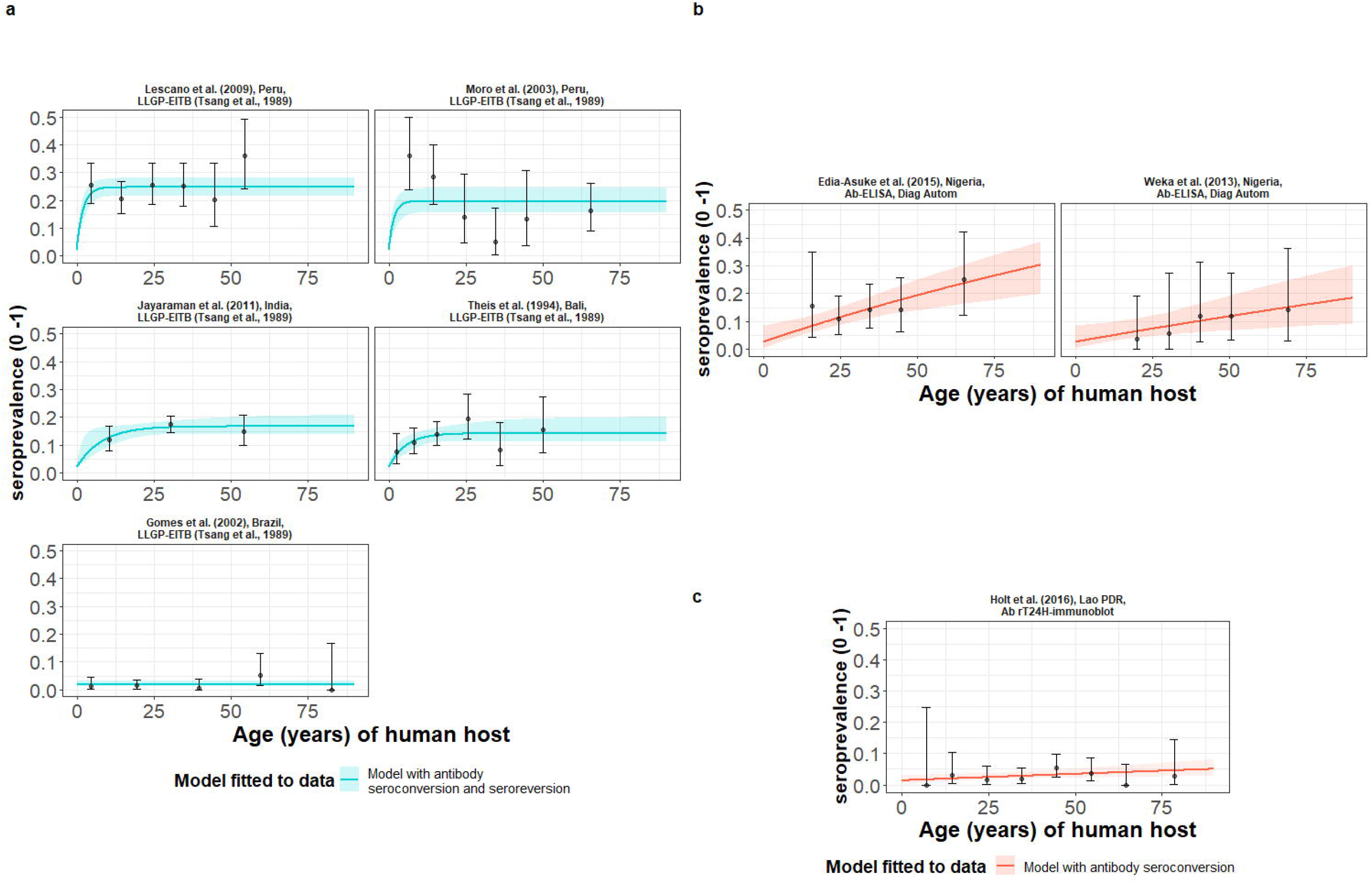
The relationship between human cysticercosis antibody (Ab)-seroprevalence and human age (in years) for each dataset. Ab-seroconversion (simple) or Ab-seroconversion with Ab- seroreversion (reversible) catalytic models (**a & b**) jointly fitted to multiple datasets (single diagnostic sensitivity and specificity values estimated; dataset-specific *λ*_*sero*_ and *ρ*_*sero*_ estimates obtained) and (**c**) models independently fitted to a single dataset, including 95% confidence intervals associated with observed Ab-seroprevalence point estimates. Bayesian Markov chain Monte Carlo methods were used to fit the models to data, with the estimated parameter posterior distributions used to construct predicted (all age) seroprevalence curves and associated 95% Bayesian credible intervals (BCIs). Best-fitting models were selected using the deviance information criterion (DIC); both models presented if difference between DIC < 2 (both models have similar support based on the data); a difference > 10 units indicates that the models are significantly different and therefore only superior fitting model (lowest DIC) is presented). The non-zero predicted seroprevalence at age 0 is due to less than 100% specificity for all tests. The 95% confidence intervals (95% CIs) for age-seroprevalence data-points are calculated by the Clopper-Pearson exact method.

### Global human cysticercosis (HCC) antigen seroprevalence

HCC infection acquisition with infection loss (reversible model) provided an improved fit for models fitted jointly to multiple datasets based on B158/B60 Ag-ELISA (Brandt et al., 1992; Dorny et al., 2000) (Table 3 and Figure 4a), found in one hyperendemic setting (all-age HCC seroprevalence of 21.7%) in the DRC (Kanobana et al., 2011), and endemic settings in Zambia (Mwape et al., 2012), Burkina Faso (Sahlu et al., 2019), Lao PDR (Conlan et al., 2012) and Cameroon (Ngeuekam et al., 2003) (all-age HCC seroprevalences from 0.7% to 5.8%). For models fitted to the single dataset from a hyperendemic setting in Kenya (6.61%) (Wardrop et al., 2015), using the HP10 Ag-ELISA (Harrison et al., 1989), the (simple) model without infection loss provided an improved fit (Table 3 and Figure 4b). Supplementary File 1: Supplementary File Table S5 presents all model fits and DIC scores.

**Table 3.**
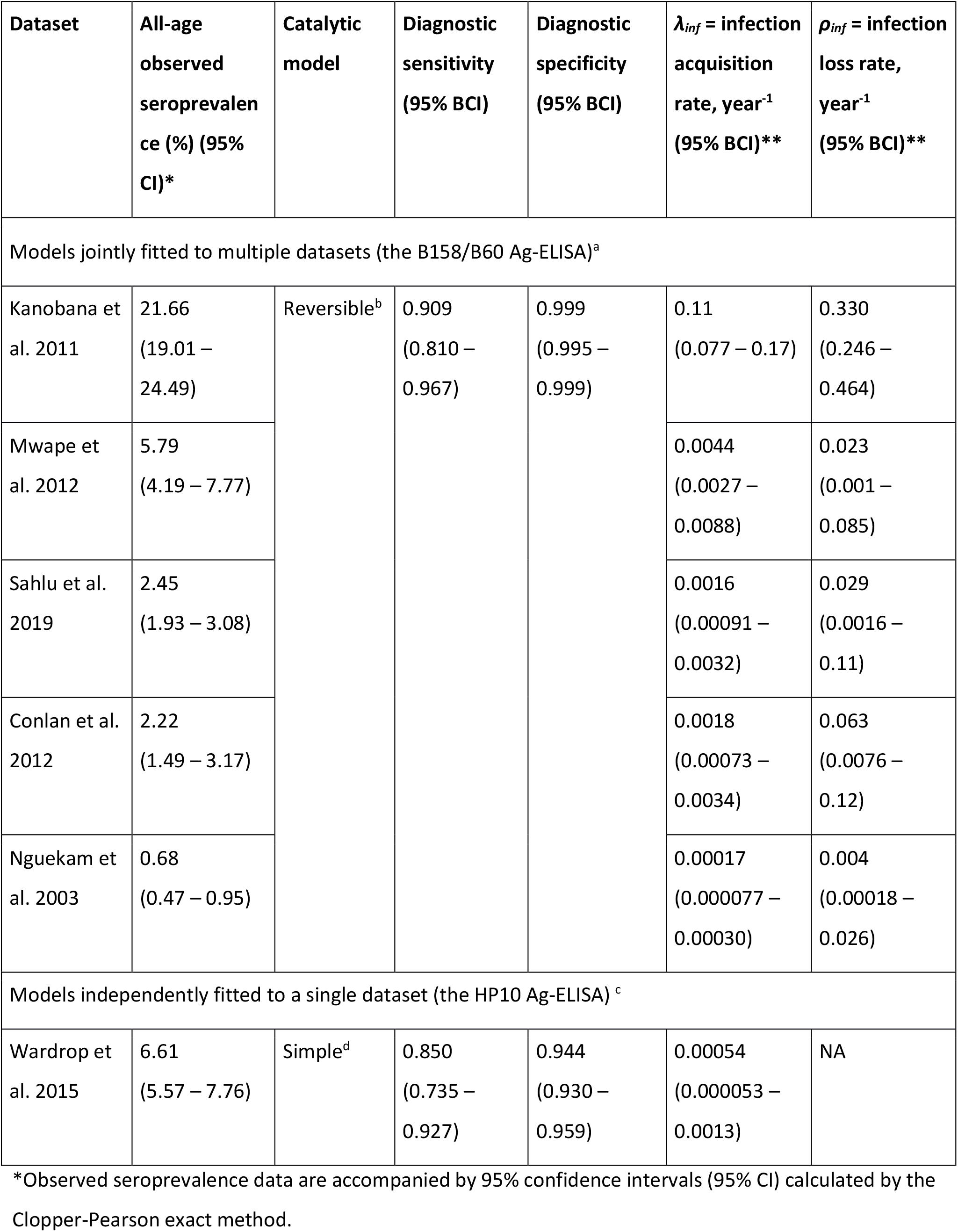
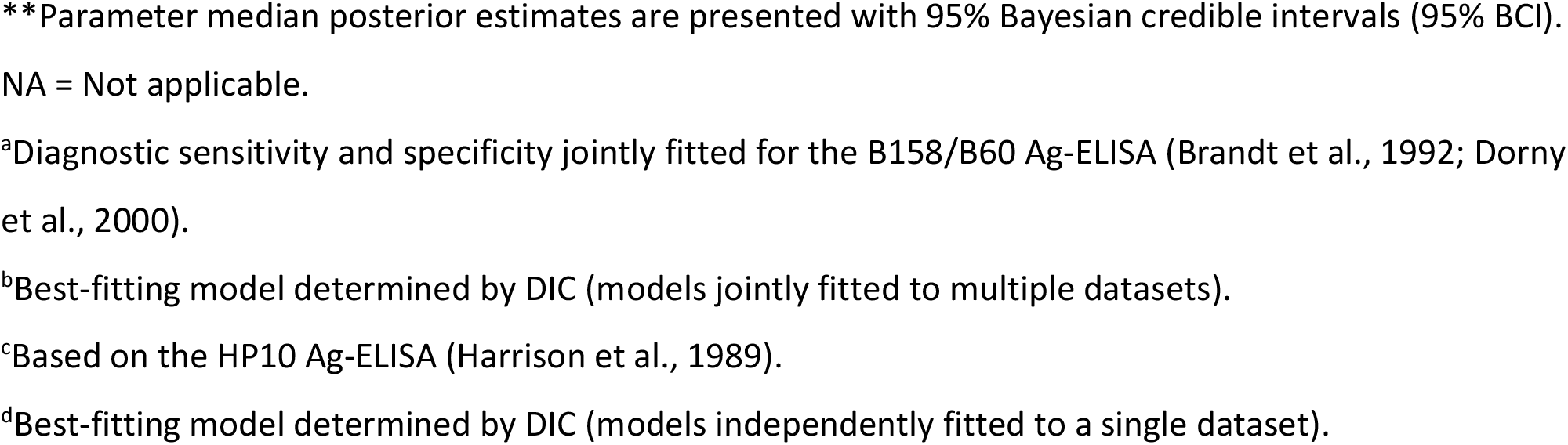
Antigen seroprevalence and parameter estimates for the best-fit catalytic models fitted to each observed human cysticercosis (antigen) age-seroprevalence dataset (ordered by decreasing all-age seroprevalence).

| Dataset | All-age<br>observed<br>seroprevalen<br>ce (%) (95%<br>CI)* | Catalytic<br>model | Diagnostic<br>sensitivity<br>(95% BCI) | Diagnostic<br>specificity<br>(95% BCI) | $\lambda_{inf}$ = infection<br>acquisition<br>rate, year<sup>-1</sup><br>(95% BCI)** | $\rho_{inf}$ = infection<br>loss rate,<br>year<sup>-1</sup><br>(95% BCI)** |
| --- | --- | --- | --- | --- | --- | --- |
| Models jointly fitted to multiple datasets (the B158/B60 Ag-ELISA)<sup>a</sup> |  |  |  |  |  |  |
| Kanobana et<br>al. 2011 | 21.66<br>(19.01 –<br>24.49) | Reversible<sup>b</sup> | 0.909<br>(0.810 –<br>0.967) | 0.999<br>(0.995 –<br>0.999) | 0.11<br>(0.077 – 0.17) | 0.330<br>(0.246 –<br>0.464) |
| Mwape et<br>al. 2012 | 5.79<br>(4.19 – 7.77) |  |  |  | 0.0044<br>(0.0027 –<br>0.0088) | 0.023<br>(0.001 –<br>0.085) |
| Sahlu et al.<br>2019 | 2.45<br>(1.93 – 3.08) |  |  |  | 0.0016<br>(0.00091 –<br>0.0032) | 0.029<br>(0.0016 –<br>0.11) |
| Conlan et al.<br>2012 | 2.22<br>(1.49 – 3.17) |  |  |  | 0.0018<br>(0.00073 –<br>0.0034) | 0.063<br>(0.0076 –<br>0.12) |
| Nguekam et<br>al. 2003 | 0.68<br>(0.47 – 0.95) |  |  |  | 0.00017<br>(0.000077 –<br>0.00030) | 0.004<br>(0.00018 –<br>0.026) |
| Models independently fitted to a single dataset (the HP10 Ag-ELISA)<sup>c</sup> |  |  |  |  |  |  |
| Wardrop et<br>al. 2015 | 6.61<br>(5.57 – 7.76) | Simple<sup>d</sup> | 0.850<br>(0.735 –<br>0.927) | 0.944<br>(0.930 –<br>0.959) | 0.00054<br>(0.000053 –<br>0.0013) | NA |

\*Observed seroprevalence data are accompanied by 95% confidence intervals (95% CI) calculated by the Clopper-Pearson exact method.
<sup>\*\*</sup>Parameter median posterior estimates are presented with 95% Bayesian credible intervals (95% BCI).
NA = Not applicable.
<sup>a</sup>Diagnostic sensitivity and specificity jointly fitted for the B158/B60 Ag-ELISA (Brandt et al., 1992; Dorny et al., 2000).
<sup>b</sup>Best-fitting model determined by DIC (models jointly fitted to multiple datasets).
<sup>c</sup>Based on the HP10 Ag-ELISA (Harrison et al., 1989).
<sup>d</sup>Best-fitting model determined by DIC (models independently fitted to a single dataset).

**Figure 4.**
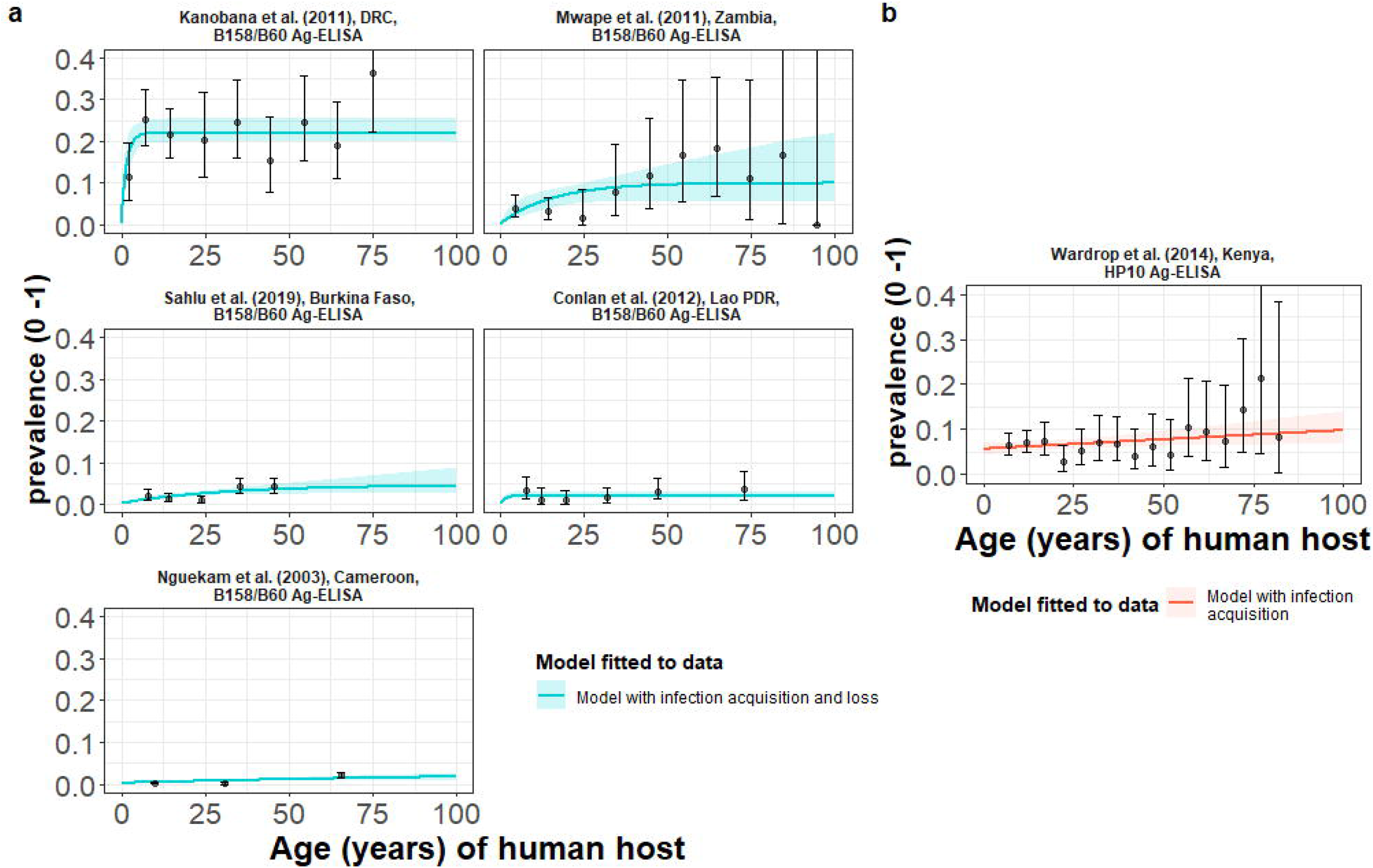
The relationship between human cysticercosis antigen (Ag)-seroprevalence and human age (in years) for each dataset. Infection acquisition (simple) or infection acquisition and loss (reversible) catalytic models (**a**) jointly fitted to multiple datasets (single diagnostic sensitivity and specificity values estimated; dataset-specific *λ*_*inf*_ and *ρ*_*inf*_ estimates obtained) and (**b**) models independently fitted to a single dataset, including 95% confidence intervals associated with observed Ag-seroprevalence point estimates. Bayesian Markov chain Monte Carlo methods were used to fit the models to data, with the parameter posterior distributions used to construct predicted (all age) seroprevalence curves and associated 95% Bayesian credible intervals (BCIs). Best-fitting model selected by deviance information criterion (DIC); both models presented if difference between DIC < 2 (both models have similar support based on the data); a difference > 10 units indicates that the models are significantly different and therefore only superior fitting model (lowest DIC) is presented). The non-zero predicted seroprevalence at age 0 is due to less than 100% specificity for all tests. The 95% confidence intervals (95% CI) for age-seroprevalence data-points are calculated by the Clopper-Pearson exact method.

### Country-wide analysis of human cysticercosis antibody seroprevalence trends in Colombia

HCC Ab-seroconversion with Ab-seroreversion (reversible model), fitted to multiple datasets using an IgG Ab-ELISA (López et al. 1988), provided an improved fit across the 23 (out of a total of 24) departments of Colombia (Flórez Sánchez et al., 2013) included in the analysis. Supplementary File 1: Supplementary Table S6 presents parameter estimates from the best-fit reversible model by department (parameter estimates for the simple model fits by department can also be found in Supplementary Table S7). Figure 5 presents the best-fit Ab-seroconversion with Ab-seroreversion (reversible) model fit to each age-seroprevalence dataset across departments (Supplementary File 1: Supplementary Figure S3 zooms into model fits in medium to lower all-age seroprevalence departments for improved resolution). One department (Bolívar) was omitted due to difficultly fitting to such a distinct age-seroprevalence profile (prevalence peak in early ages).

**Figure 5.**
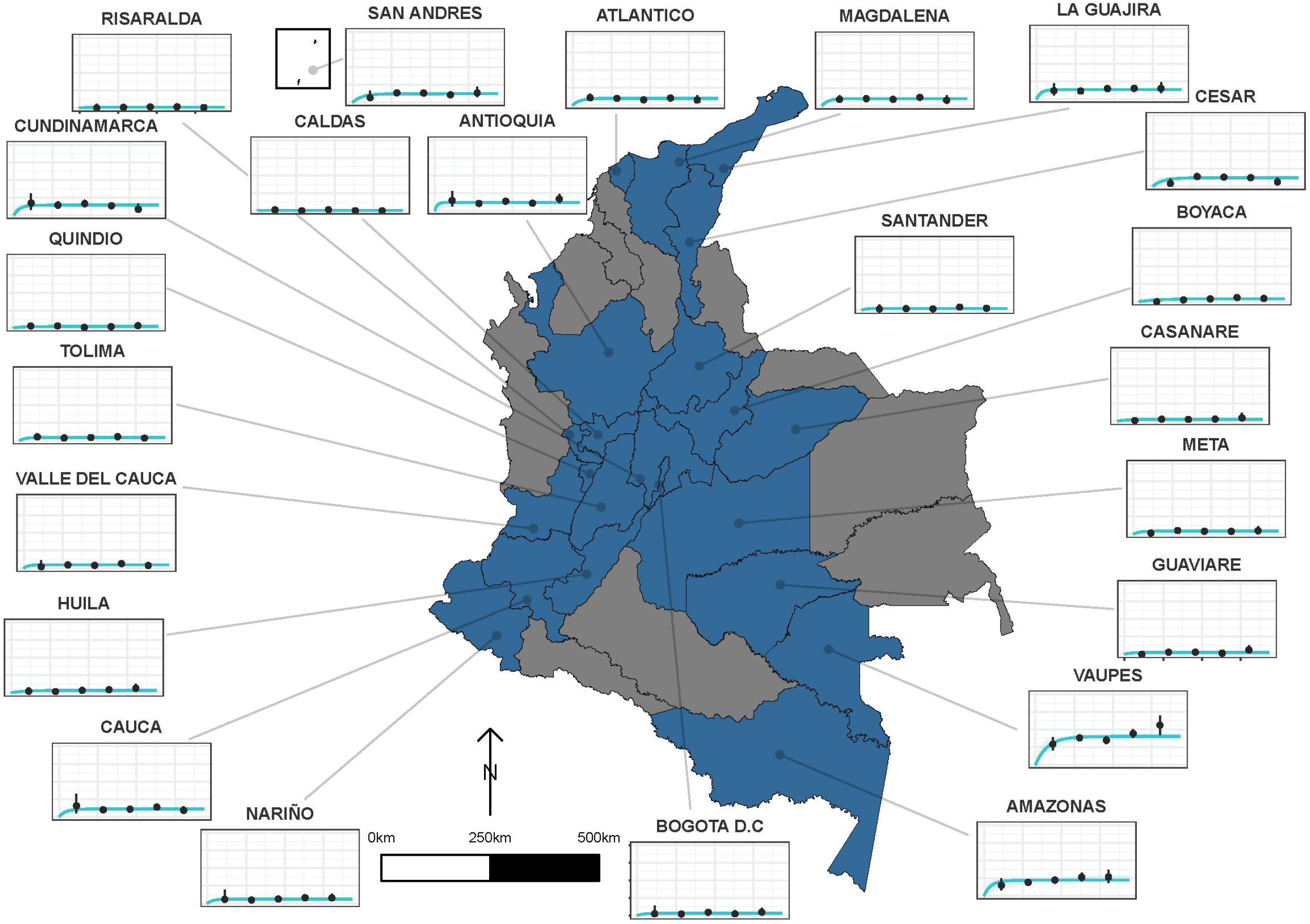
The relationship between human cysticercosis antibody (Ab)-seroprevalence and human age (in years) for 23 departments in Colombia. Each graph shows observed human cysticercosis Ab age-seroprevalence data (black points) and fitted reversible model (Ab-seroconversion with Ab- seroreversion; best-fitting model). Y-axis units are from 0-1 (HCC Ab-seroprevalence), with major y-axis gridlines at 0, 0.2, 0.4, and 0.6 seroprevalence. X-axis units are from 0-80 years (human age), with major x-axis gridlines at 0, 20, 40, 60 and 80 years of age. *Note: Supplementary File 1: Supplementary Figure S7 zooms into predicted plots for a) medium all-age HCC Ab-seroprevalence (6*.*33–14*.*37% all-age seroprevalence) and b) low all-age HCC Ab-seroprevalence (0*.*48–4*.*92% all-age seroprevalence), to view more closely the age-seroprevalence trends and uncertainty around fitted seroprevalence*.

Figure 6 highlights the geographical variation in a) HCC Ab-seroconversion or FoI, b) HCC Ab- seroreversion rate and c) the HCC antibody all-age seroprevalence, across the 23 departments. In addition, Figure 6 presents substantial geographical variation in risk factors, including: d) the proportion of individuals owning pigs, and e) the proportion of individuals reporting open defecation practices by department (Supplementary File 1: Supplementary Figure S4 highlights variation in the proportion of pigs being kept under free-ranging management practices and the proportion of pigs being kept under free-ranging/mixed practices in those owning pigs (*n*=3,157)).

**Figure 6.**
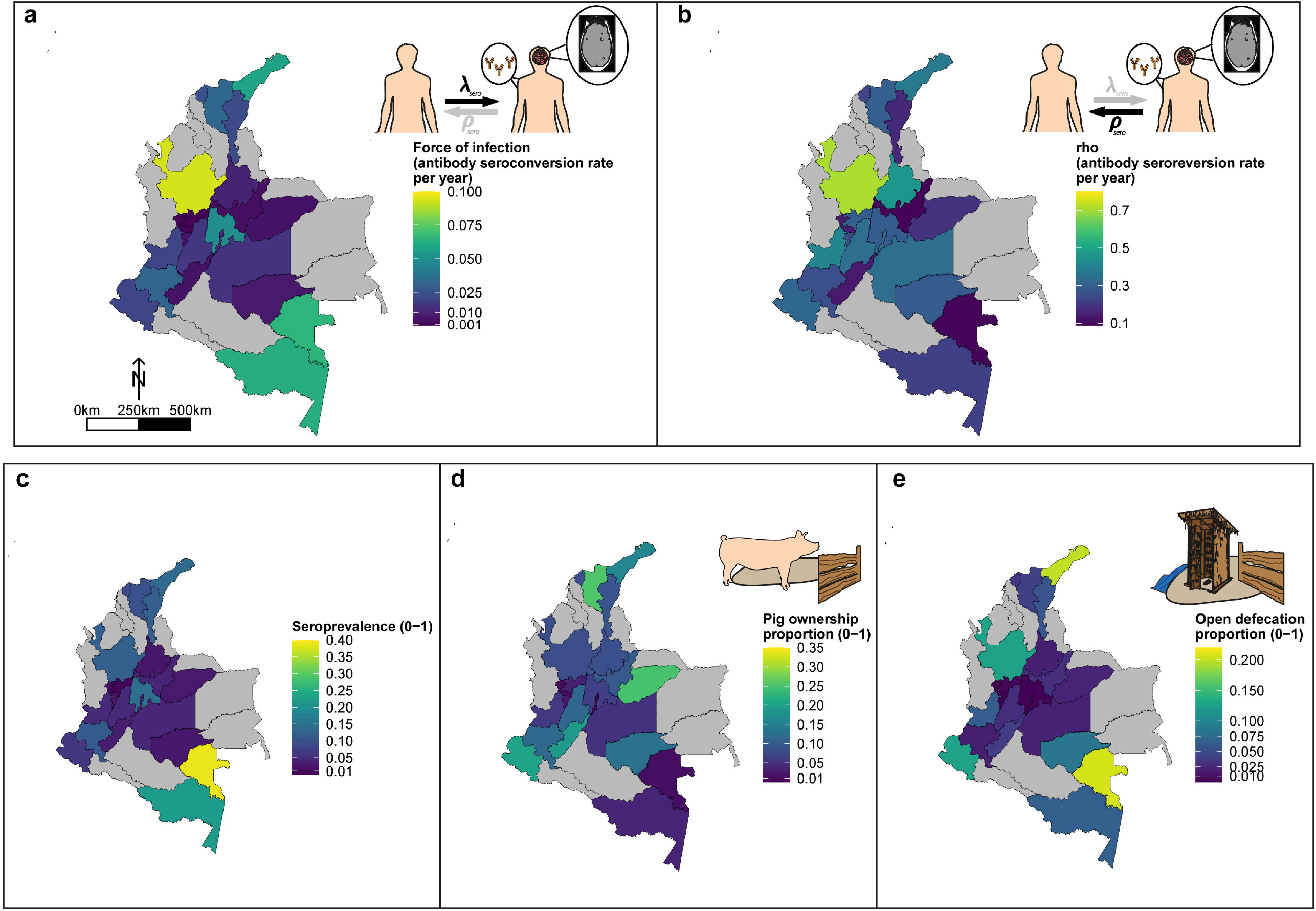
Geographic variation in, a) HCC antibody (Ab)*-*seroconversion rate (*λ*_sero_ or FoI), and b) HCC Ab*-*seroreversion rate (*ρ*_sero_), c) human cysticercosis Ab all-age seroprevalence data, d) pig ownership proportion and e) open defecation reported proportion by department. The FoI (*λ*_sero_) and Ab*-* seroreversion (*ρ*_sero_) rates are parameter estimates obtained from the best-fit model with HCC Ab*-* seroconversion and Ab*-*seroreversion (reversible model). Note that San Andrés department is not clearly shown because of its size (small islands located in the top-left of a-c maps). For context, an all-age HCC Ab*-*seroprevalence = 0.126, HCC Ab*-*seroconversion rate (FoI or *λ*_sero_) = 0.023 year-1, and HCC *Ab-*seroreversion rate (*ρ*_sero_) = 0.19 year-1 was obtained in San Andrés

### Force-of-infection across settings

A more intuitive approach to understanding the FoI (*λ*) is to consider its reciprocal, which here corresponds to the average time until humans become Ab-seropositive (1/*λ*_*sero*_) or infected (1/*λ*_*inf*_) as inferred through Ag-seropositivity. Equally, the reciprocal of *ρ* relates to the average duration that humans remain Ab-seropositive (1/*ρ*_*sero*_) or infected (1/*ρ*_*inf*_). Given the large number of estimates obtained for HCC, parameter estimates from best-fit models were compared across settings (by all-age (sero)prevalence of each dataset and by country). Figure 7 shows an overall decline in the average time (in years) until humans become HCC Ab-seropositive or infected with increasing all-age HCC (sero)prevalence, noting 18 studies identified in endemic settings (0.48– 5.71% all-age HCC (sero)prevalence), and 19 estimates in hyperendemic settings (6.33–38.68% all-age HCC seroprevalence). Within countries, there was significant variation in times until humans become HCC Ab-seropositive or infected (Supplementary File 1: Supplementary Figure S5a).

**Figure 7.**
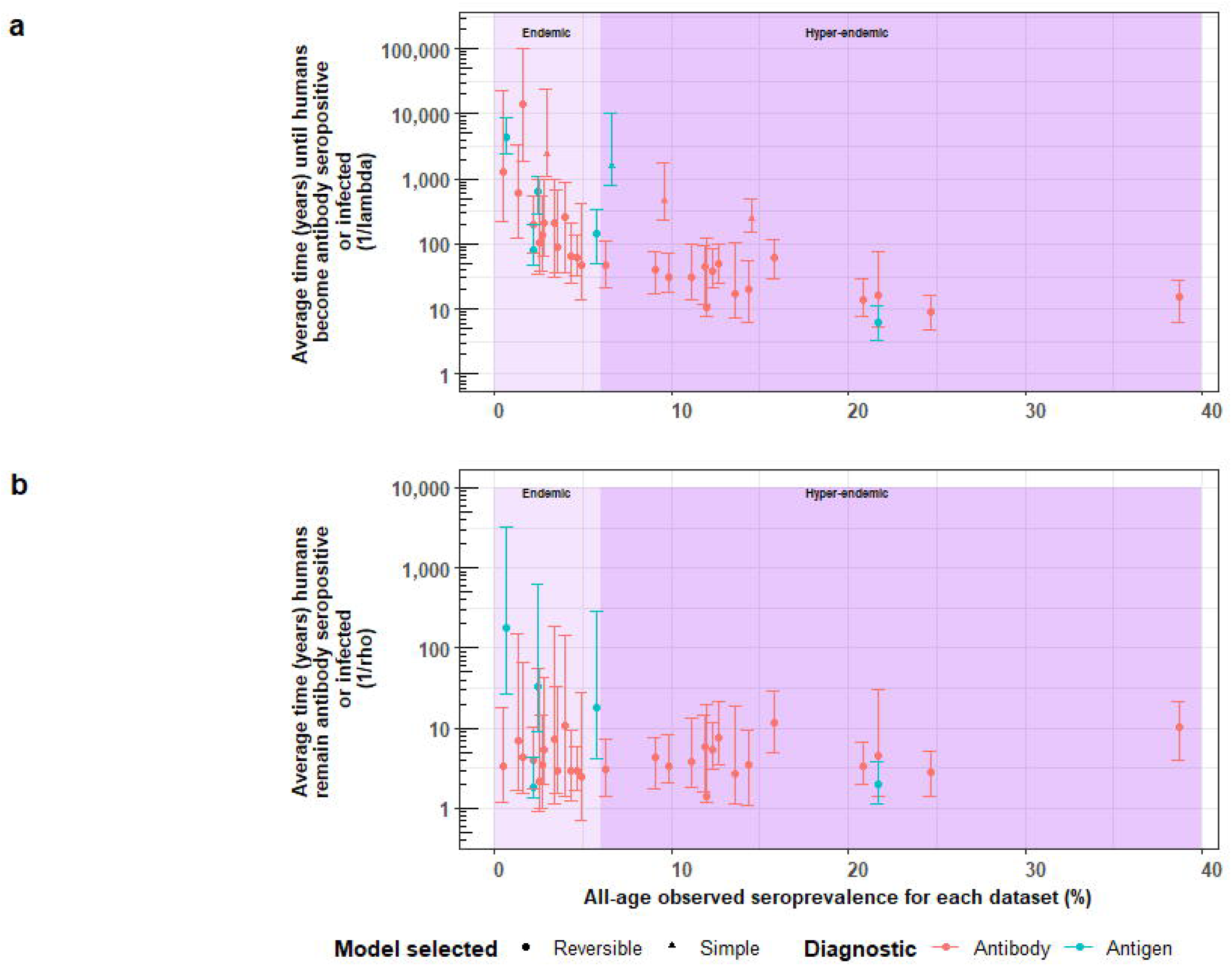
The relationship between (a) the average time until humans become cysticercosis antibody (Ab)-seropositive or infected (1/*λ*) and overall (all-age) seroprevalence, and (b) the average time humans remain cysticercosis Ab seropositive or infected (1/*ρ*) and overall (all-age) seroprevalence. The plot is stratified by proposed endemicity levels defined as endemic (>0% and <6% all-age HCC seroprevalence), and hyperendemic (≥6% all-age HCC seroprevalence).

Figure 7b presents strong evidence for similar average durations of remaining Ab-seropositive (1/*ρ*_*sero*_) across different endemicity settings and countries (Supplementary File 1: Supplementary Figure S5b). However, there was evidence for a trend of decreasing duration of humans remaining infected (1/*ρ*_*inf*_) with increasing all-age prevalence (antigen-based studies; n=5).

## Discussion

This paper presents the first global FoI estimates for *T. solium* HTT and HCC, identifying geographical heterogeneity that supports calls to implement both locally-adapted control efforts (Johansen et al., 2017; Gabriël et al., 2017), and setting-specific parameterisations of *T. solium* transmission models to support such efforts (Dixon et al., 2019). We have also placed our HTT FoI estimates in the context of (putative) endemic or hyperendemic settings, with 0.44 per 1,000 people per year for a (single) endemic setting, and from 9.6 to 21 per 1,000 per year for 2 hyperendemic settings. HCC FoI estimates ranged from 0.086 to 21 per 1,000 per year in (18, including Colombian departments) endemic settings (0.086–21 for Ab-based surveys; 0.17–4.4 for Ag-based surveys), and from 0.54–120 per 1,000 per year in (19) hyperendemic settings (2.3–120 for Ab-based surveys; 0.54–110 for Ag-based surveys). Further work will be required to refine the proposed (and preliminary) characterisation of endemicity levels, perhaps linked to severity/morbidity (as in other NTDs (Prost et al., 1979; Smith et al., 2013). This will be relevant to inform what constitutes a hyperendemic setting in need of intensified control interventions, as proposed in the WHO 2021– 2030 goals for *T. solium* (World Health Organization, 2020). Our work also follows the 5 principles distilled by Behrend et al. (2020) for communicating policy-relevant NTD modelling to stakeholders and implementation partners (Supplementary File 1: Supplementary File Table S8).

Across epidemiological settings, there was a clear trend of decreasing average time until humans become HCC Ab-seropositive or infected as inferred through Ag-seropositivity (the reciprocal of *λ*_*sero*_ *or λ*_*inf*_) with increasing all-age seroprevalence. This makes intuitive sense as the FoI increases in settings with more intense transmission (reflected by a higher all-age seroprevalence). It was not possible to discern whether such trends exist for HTT given the limited availability of HTT datasets with age-stratified prevalence data. Infection loss with infection acquisition was indicated for two HTT copro-Ag-ELISA-based surveys, and five of six HCC antigen-based surveys, supporting inclusion of parameters representing recovery from HTT and HCC in *T. solium* transmission models (Dixon et al., 2019). Our infection loss rates for HTT indicate that the duration of adult tapeworm infection (the reciprocal of *ρ*_*inf*_) ranges from 1.3 to 1.4 years (with uncertainty bounds from 1.0 to 2.8 years), aligning with literature estimates suggesting that adult tapeworms live for less than 5 years (Garcia et al., 2014), although our values are only based on two studies. HCC Ab-seroreversion rates (or infection loss rates) from this analysis are also consistently larger than HCC Ab-seroconversion rates (or infection acquisition rates), in agreement with comparisons of HCC Ab-seroreversion to Ab- seroconversion (or Ag-seroreversion to Ag-seroconversion) estimates in the literature, which are generally based on cumulative seroconversion and seroreversion proportions, or rule-based modelling approaches (Dermauw et al., 2018; Garcia et al., 2001; Coral-Almeida et al., 2015; Mwape et al., 2013).

Some of the modelled estimates presented here indicate large differences between FoI and Ab- seroreversion/infection loss rates. These differences are particularly evident in datasets, such as Brazil (Gomes et al., 2002) and Risaralda Department in Colombia (Flórez Sánchez et al., 2013), where the reversible model was selected for flat age-(sero)prevalence profiles in low (sero)prevalence settings, where the seroreversion parameter provided minimal information with large uncertainty. The flat age-(sero)prevalence in these settings may also be likely explained by false-positives generated given the slightly sub-optimal performance of the serological antibody diagnostic used (e.g. posterior median specificity estimates of 98% in Brazil (Gomes et al., 2002) for the antibody LLGP-EITB test (Tsang et al., 1989), highlighting issues with imperfect specificity of diagnostics in settings with very low transmission intensity.

The high cysticercosis Ab-seroreversion rates indicated across datasets are likely indicative of substantial transient responses generated from exposure without establishment of infection. This may also explain the high all-age Ab-seroprevalence estimates observed in field studies (Garcia et al., 2001). Even with Ag-based surveys, a marker of infection rather than exposure, Ag-positivity may still indicate some degree of transient responses due to partial establishment of (immature) cysts or establishment followed by rapid degeneration of (mature) cysts (Mwape et al., 2012). One expectation was that Ab-seroreversion and infection loss rates are fundamentally biological processes so would be fairly consistent across settings. Since Ab-seroreversion or infection loss parameters were allowed to vary among settings, the consistency of the posterior estimates for these parameters indicates that rates of HCC Ab-seroreversion are stable (i.e., no relationship between all-age Ab-seroprevalence and the average duration humans remain Ab-seropositive, Figure 7b; no relationship between the Ab-seroconversion and Ab-seroreversion rates, Supplementary Figure S6), confirming this *a priori* expectation. The average duration humans remain infected (i.e., the reciprocal of the HCC infection loss rate), however, decreases with increasing seroprevalence (and a positive relationship between infection acquisition rates and infection loss rates, Supplementary Figure S6). Such a signal seen for Ag-based estimates, albeit with only 5 observations, may indicate increased transient responses in higher transmission settings due to more partial establishment of infection. There is however an absence of wider literature to support the hypothesis of exposure intensity affecting probability of successful parasite establishment, but such (transmission intensity-dependent) parasite establishment processed have been incorporated into transmission models for other NTDs (Hamley et al., 2019), highlighting an area of future research with implications for control efforts. In the case of Ab-seroreversion rates, where more data are available compared to infection loss rates, it may be suitable in future work to jointly fit the *ρ*_*sero*_ parameter for reversible models across settings.

This analysis also provides, for the first time, a detailed country-level analysis, focused on Colombia, exploring the geographical variation of key epidemiological metrics (FoI and antibody seroreversion) alongside understanding potential drivers of these trends, using a rich dataset generated by Flórez Sánchez and colleagues (2013). Our results identify substantial subnational variation in HCC FoI (measured by Ab-seroconversion) and Ab-seroreversion rates. There appears to be only a limited relationship between these epidemiological quantities, Ab-seroprevalence and other risk factors, including open defecation and pig ownership at the department level, highlighting the complex nature of exposure drivers for HCC. For example, the southeastern rural departments of Vaupés and Amazonas show the highest all-age seroprevalence and FoI estimates. But while Vaupés has one of the highest reported proportions of open defecation, pig ownership is low in both departments. Notwithstanding, substantial subnational variation indicates the need for tailored subnational approaches to control. Spatial variation has been explored formally for this dataset by including spatial structure (i.e., spatial correlation in seropositivity estimated up to approximately 140 km at the Municipality level) into a risk factor analysis, and identifying hot spots of unexplained residual clustering in northern and southern municipalities in Colombia (Galipó et al., 2021). The findings from this study further supports the requirement for designing subnational and targeted interventions (risk groups such as those with low education levels and displaced persons), alongside the need to understand epidemiological dynamics at the subnational level.

A potential limitation of this study is the assumption that diagnostic sensitivity and specificity for a single diagnostic test do not vary substantially among settings (implied by estimating a single posterior for specificity and sensitivity using data from multiple surveys that used the same diagnostic). In reality, there may be some variation in diagnostic performance between settings, particularly for HTT serological diagnostics. For example, the widely used copro-Ag ELISA for detecting adult tapeworm infection is not species specific (Ng-Nguyen et al., 2017). Literature estimates indicate high specificity of HCC diagnostics; however, a proportion of positive results by both HCC Ag- and Ab- diagnostics may be due to transient responses and cross-reactions following exposure to the eggs of other other helminths including *Taenia saginata, Echinoccous granulosus* and *Schistosomia species* (Lightowlers et al., 2016b; Noh et al., 2014; Kojic et al., 2003; Furrows et al., 2006). Limited information exists on the co-distribution and prevalence of potentially cross-reactive *Taenia* species (such as *Taenia saginata*) and other helminths to determine the relative contribution of within- and between-location variability in the performance of specific diagnostic tests.

The prevailing (and most parsimonious) assumption governing the behaviour of the catalytic models fitted in this analysis is of a constant FoI with respect to age. Collated age-(sero)prevalence profiles presented here largely reveal increasing (sero)prevalence with age, or saturation with age through Ab-seroreversion/infection loss, but this behaviour can also be observed with age-dependent infection rates (Grenfell and Anderson, 1985). However, the available data limit the testing of more complex FoI functions. A few datasets appeared indeed to deviate from the basic assumption, for example for HTT dynamics in the DRC (Madinga et al., 2017) and Bolívar in Colombia (Flórez Sánchez et al., 2013). While it was not possible to fit the current formulation of catalytic models to these datasets, the specific age-prevalence peaks in younger ages may indicate other drivers such as heterogeneity in exposure (by age) or age-related immunity mechanisms. The observations of HTT copro-Ag-ELISA age-prevalence peaks/odds of infection in children from Peru, Guatemala and Zambia (Garcia et al., 2003a; Allan et al., 1996; Mwape et al., 2015) suggest that these trends might be more widespread. Age peaks in HTT prevalence could indicate the need for targeted mass drug administration (MDA) programmes, such as delivery of anthelminthics (praziquantel) to school-age children, an approach that could be integrated into existing schistosomiasis control programmes where HTT and schistosomiasis are co-endemic. Should more age-prevalence data become available, especially for HTT, fitting models with age-varying FoI may become relevant as indicated in other NTDs (Gambhir et al., 2009).

The results presented here, particularly for HCC, where many datasets were available, suggest marked geographical heterogeneity in FoI and associated all-age (sero)prevalence estimates. This indicates substantial variation in the intensity of *T. solium* transmission among endemic settings. There is strong evidence for both HCC Ab-seroreversion and infection loss, likely due to transient dynamics which highlights the need for careful interpretation of cross-sectional (sero)prevalence survey estimates. Catalytic models provide useful tools for interpreting such data, which are far more abundant than a few longitudinal surveys reported in the literature. We also quantify, for the first time, the presence of substantial subnational variation in exposure to HCC, highlighting the need for tailored, sub-national control strategies. More work is also required to understand whether age-prevalence peaks (as observed for HTT) are more commonplace, and whether age-targeted control strategies may be required under specific epidemiological and socioeconomic conditions.

## Materials and methods

### Identifying relevant literature, data sources and data extraction

Published articles with HTT and HCC age-(sero)prevalence *or* age-infection data suitable for constructing age-stratified profiles were identified through two routes. Firstly, by extracting eligible studies from a systematic review of *T. solium* HTT and HCC (sero)prevalence global ranges (Coral-Almeida et al. (2015)), and secondly, by updating the Coral-Almeida et al. (2015) review, using the same strategy (see Supplementary File 1: Supplementary Text S1), for the period 01/11/2014 to 02/10/2019.

### Sub-national dataset for Colombia

Flórez Sánchez et al. (2013) conducted a country-level survey of HCC antibody seroprevalence across 24 of Colombia’s 32 administrative departments, sampling 29,360 individuals. Permission was granted from *Instituto National de Salud* to analyse these data to construct age-seroprevalence profiles. The study collected human cysticercosis Ab-seroprevalence data in the period 2008–2010, alongside risk-factor data using a three-stage clustered random sampling framework. To maintain suitable sample sizes for model fitting, age-seroprevalence profiles were constructed at the department level (*n* = 850–1,291; Supplementary File 1: Supplementary Figure S7), rather than at municipality level (*n* = 40–1,140). Age-seroprevalence profiles stratified by sex in each department revealed no clear differences (Supplementary File 1: Supplementary Figure S8a), therefore age-seroprevalence data were not stratified by sex for further analysis (but see Galipó et al., 2021). Due to difficulty fitting the specific catalytic models to data from the department of Bolívar, only 23 of 24 departments were assessed (*n* = 28,100).

### Force-of-infection modelling for human taeniasis and human cysticercosis

The FoI, which describes the average (per capita) rate at which susceptible individuals seroconvert (become Ab-positive) or become infected, was estimated for HTT and HCC for each dataset. Simple and reversible catalytic models (Figure 1), originally proposed for fitting to epidemiological data by Muench (1934), were fitted to HTT and HCC age-(sero)prevalence profiles. Model parameters fitted to Ab-datasets were interpreted as Ab-seroconversion (*λ*_*sero*_) and Ab-seroreversion (*ρ*_*sero*_) rates, while (copro-) Ag-datasets were interpreted as infection acquisition (*λ*_*inf*_) and infection loss (*ρ*_*inf*_) rates (Figure 1 provides further details of model structure and parameter interpretation). Like the FoI, the infection loss/ seroreversion parameter (*ρ*) was also permitted to vary across settings (Dermauw et al., 2018).

The true prevalence *p*(*a*) at age *a* is given for the simple model by,

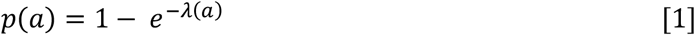

and for the reversible model by,

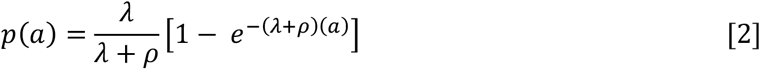

### Model fitting and comparison

Analyses were performed in R version 4.0.5 (2021), following Dixon et al. (2020). A likelihood was constructed assuming that the observed data (representing a binary presence/absence of markers related to exposure or infection) are a realization of an underlying binomial distribution with probability p(a), given by the catalytic model, and adjusted to give the observed prevalence, p’(a), by the sensitivity (se) and specificity (sp) of the diagnostic adopted in the respective datasets (Diggle, 2011),

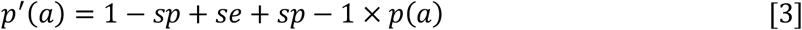

Therefore, the likelihood of the data on the number of observed Ab-seropositive or infected humans of age a, r(a), from n(a) humans is,

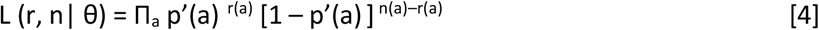

where θ denotes, generically, the catalytic model (i.e. FoI, seroreversion/infection loss) and diagnostic performance (i.e., sensitivity and specificity) parameters. When the same diagnostic was used across surveys, sensitivity and specificity were assumed to be the same among surveys, yielding a single posterior distribution for the diagnostic performance (sensitivity, specificity) for each test (whilst retaining dataset-specific estimates of λ and ρ). The approach captures uncertainty in diagnostic sensitivity and specificity, but does not permit variation in performance across settings.

A Bayesian Markov chain Monte Carlo (MCMC) Metropolis–Hastings sampling algorithm was implemented to estimate the parameter posterior distribution f(θ|r, n), assuming uniform prior distributions for λ and ρ (Table 4). A weakly informative prior for λ, assuming a lognormal distribution (mean informed by the median estimate from the simple model fit, and a standard deviation of 1), was used for reversible model fits to prevent λ chains drifting to flat posterior space and failing to converge.

**Table 4.** Range for force-of-infection (FoI), *λ*, and seroreversion / infection loss, *ρ*, uniform priors.

| *T. solium*<br>indicator | $\lambda$: limits on<br>uniform prior<br>(year<sup>-1</sup>) | $\rho$: limits on<br>uniform prior<br>(year<sup>-1</sup>) | Rationale |
| --- | --- | --- | --- |
| HTT (copro-antigen<br>(Ag) and<br>antibody<br>(Ab)) | Minimum: 0<br>Maximum: 12 | Minimum: 0.333<br>Maximum: 1 | $\lambda$: maximum seroconversion rate of 12 year<sup>-1</sup> = 1 month<sup>-1</sup> ($1/\lambda$), representing the lower limit before humans become copro-Ag positive (i.e., infected) or Ab-seropositive of 1 month<br>$\rho$: minimum Ab-seroreversion or infection loss rate of 0.333 year<sup>-1</sup> = 0.0277 month<sup>-1</sup>, or 36-month duration ($1/\rho$) for humans to remain copro-Ag (i.e., infected) or Ab- seropositive, reflecting the upper average limit assumed for the life- expectancy of adult *Taenia solium* of 3 years (Gonzalez et al., 2002); a maximum of 0.0833 month<sup>-1</sup> represents the minimum time humans remain copro-Ag (i.e., infected) or Ab-seropositive reflecting the minimum life-expectancy of the adult worm of 12 months (Garcia et al., 2003b) ($1/\rho$) |
| HCC<br>(antigen<br>and<br>antibody) | Minimum: 0<br>Maximum: 12 | Minimum: 0<br>Maximum: 12 | Both $\lambda$ and $\rho$: maximum infection acquisition/loss or Ab-seroconversion / Ab-seroreversion rate of 12 year<sup>-1</sup> = 1 month<sup>-1</sup> represents the lower limit before humans acquire infection or become Ab-seropositive ($1/\lambda$), or remain HCC antigen (i.e., infected) or Ab-seropositivity ($1/\rho$) of 1 month. |
| *N.B.* The reciprocal of the rates of $\lambda$ and $\rho$ give the duration of susceptibility ($1/\lambda$; $1/\lambda_{sero}$ for Ab-based, or $1/\lambda_{inf}$ for Ag-based) or of remaining infected or Ab-seropositive ($1/\rho$; $1/\rho_{sero}$ for Ab-based, or $1/\rho_{inf}$ for Ag-based). HTT: human taeniasis; Ab: antibody; Ag: antigen; HCC: human cysticercosis |  |  |  |

Informative beta distribution priors for the diagnostic sensitivity and specificity were defined to capture literature estimates of the mean and 95%CIs for these parameters; Supplementary File 1: Supplementary Table S1, Supplementary Figure S9 and S10 provide more detail).

A maximum of 20,000,000 iterations were run for models fitted simultaneously to multiple datasets (2,000,000 for models fitted to single datasets) to obtain a sufficient sample to reduce autocorrelation through substantial subsampling, with the first 25% of runs being discarded as burn-in. The parameter posterior distributions, used to generate fitted prevalence curves and associated uncertainties for each model fit, were summarised using the median and 95% Bayesian credible intervals (95% BCIs). Simple and reversible model fits were compared using the deviance information criterion (DIC) (Spiegelhalter et al., 2002), with the model producing the smallest DIC score being selected.

## Data Availability

All age-(sero)prevalence data are available in the following data repository: http://doi.org/10.14469/hpc/10047. Original data for two datasets available (under the Creative Commons Attribution License; CC BY 4.0) from the International Livestock Research Institute open-access repository (http://data.ilri.org/portal/dataset/ecozd) referenced in Holt et al. (2016) and University of Liverpool open-access repository (http://dx.doi.org/10.17638/datacat.liverpool.ac.uk/352) referenced in Fevre et al. (2017).

http://data.ilri.org/portal/dataset/ecozd

## Acknowledgements

NA

## Data and code availability

Code to replicate the analysis is provided at https://github.com/mrc-ide/human_tsol_FoI_modelling.

All age-(sero)prevalence data are available in the following data repository: http://doi.org/10.14469/hpc/10047. Original data for two datasets available (under the [Creative Commons Attribution License; CC BY 4.0](https://creativecommons.org/licenses/by/4.0/)) from the International Livestock Research Institute open- access repository (http://data.ilri.org/portal/dataset/ecozd) referenced in Holt et al. (2016) and University of Liverpool open-access repository (http://datacat.liverpool.ac.uk/352/) referenced in Fèvre et al. (2017).

