## Supplementary File 1 for "Global Force-of-Infection Trends for Human *Taenia solium* Taeniasis/Cysticercosis"

### Supplementary Figures

#### PRISMA 2009 Flow Diagram

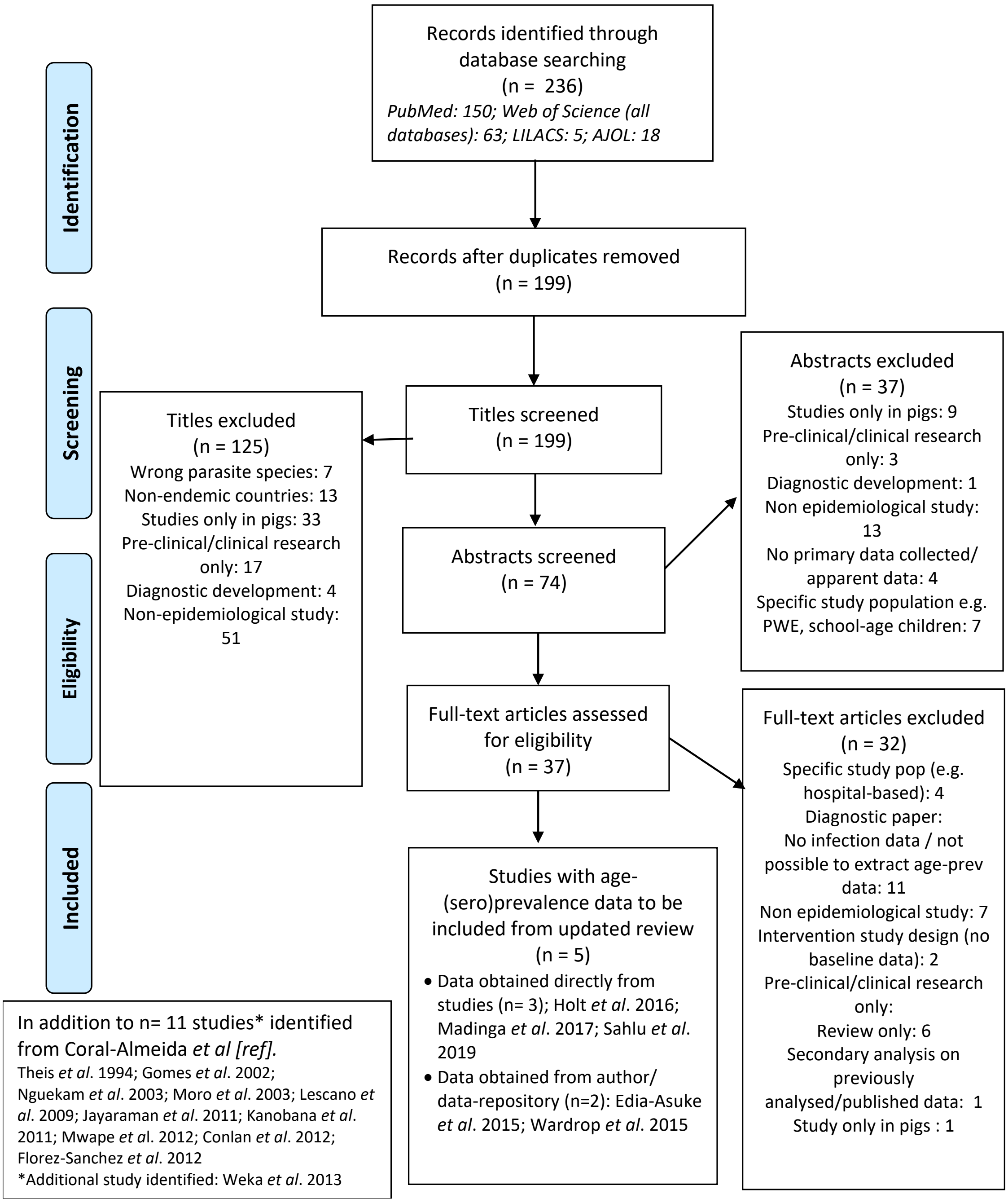


**Supplementary Figure S1. Published articles on age-(sero)prevalence profiles identified in the updated literature search using a PRISMA (2015) search.** Additional studies identified from the Coral-Almeida et al. (2015) review also shown. LILAC: Latin American and Caribbean Health Sciences Literature; AJOL: African Journals Online

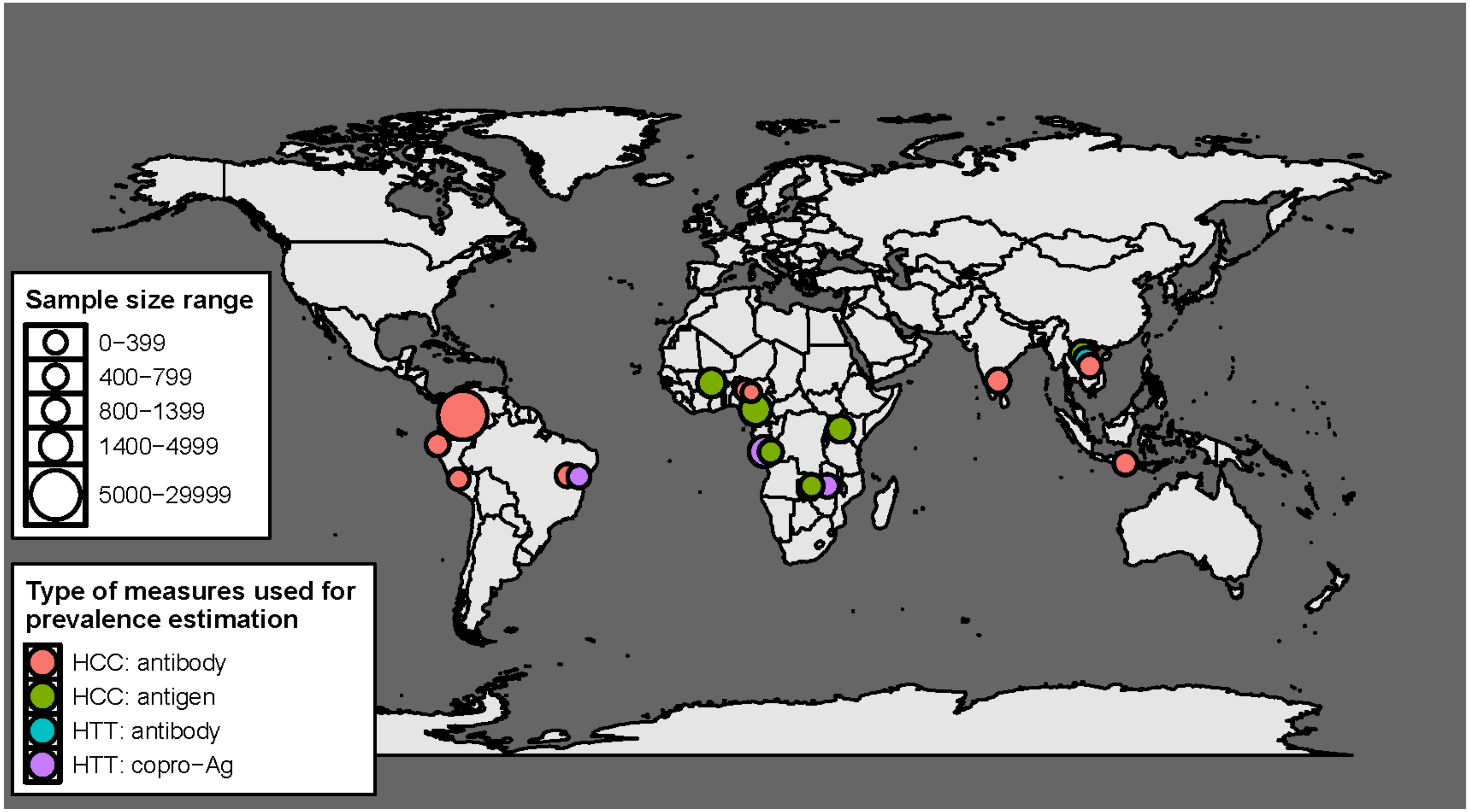


**Supplementary Figure S2. Geographical distribution of studies with human taeniasis (HTT) and human cysticercosis (HCC) age-(sero) prevalence data included in the final analysis (n = 16) by diagnostic target.** There are 19 indicators in total, as three studies provided more than one indicator (Gomes et al. (2002); Mwape et al. (2012); Holt et al. (2016)).

a) medium all-age seroprevalence Departments (6.33-14.37%)

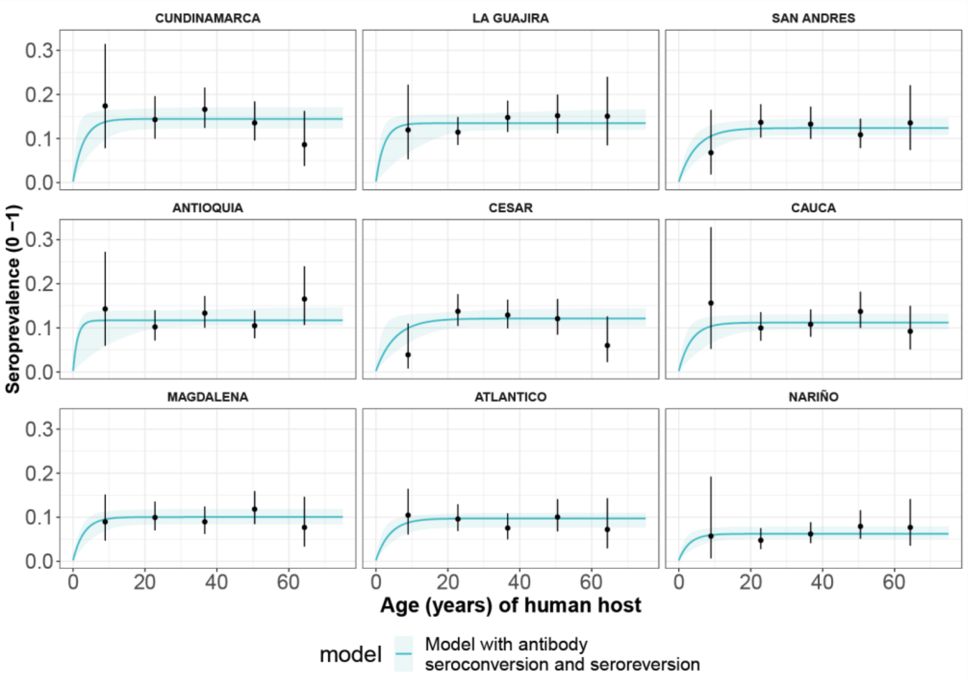


b) low all-age seroprevalence Departments (0.48-4.92%)

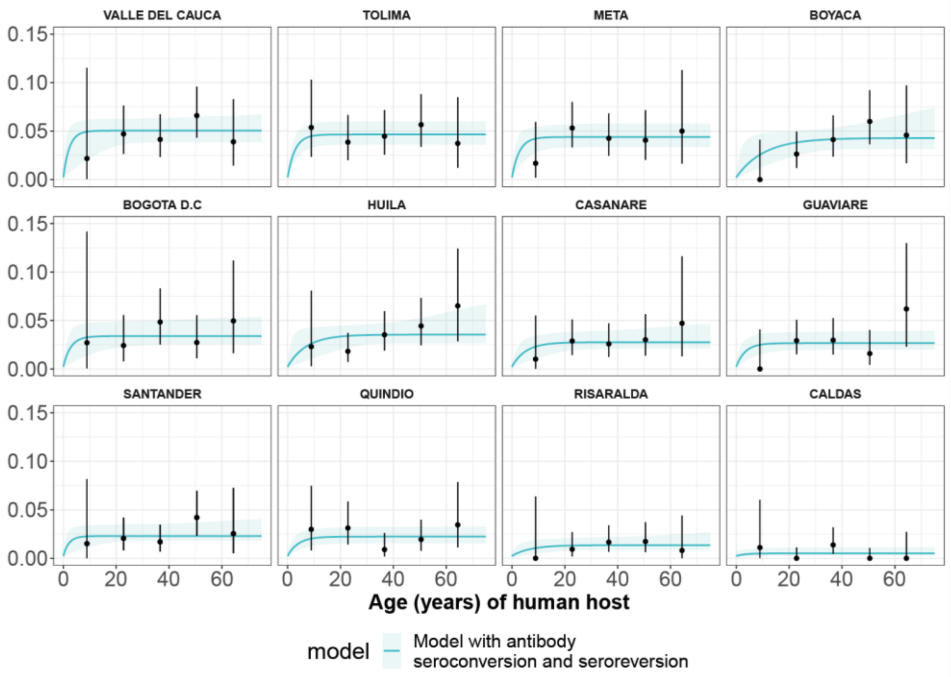


**Supplementary Figure S3. Human cysticercosis antibody (Ab) age-seroprevalence profiles and (selected by Deviance Information Criterion) model with seroconversion and seroreversion model fits to (a) medium all-age Ab-seroprevalence Colombian Departments (6.33-14.37%) and (b) low all-age Ab-seroprevalence Colombian Departments (0.48-4.92%)**. For (a) and (b), Departments are arranged in order of highest to lowest overall Department Ab-seroprevalence. Note the different y-axis scale for a) medium all-age Ab-seroprevalence Departments (0-0.3) and b) low all-age Ab-seroprevalence Departments (0-0.15).

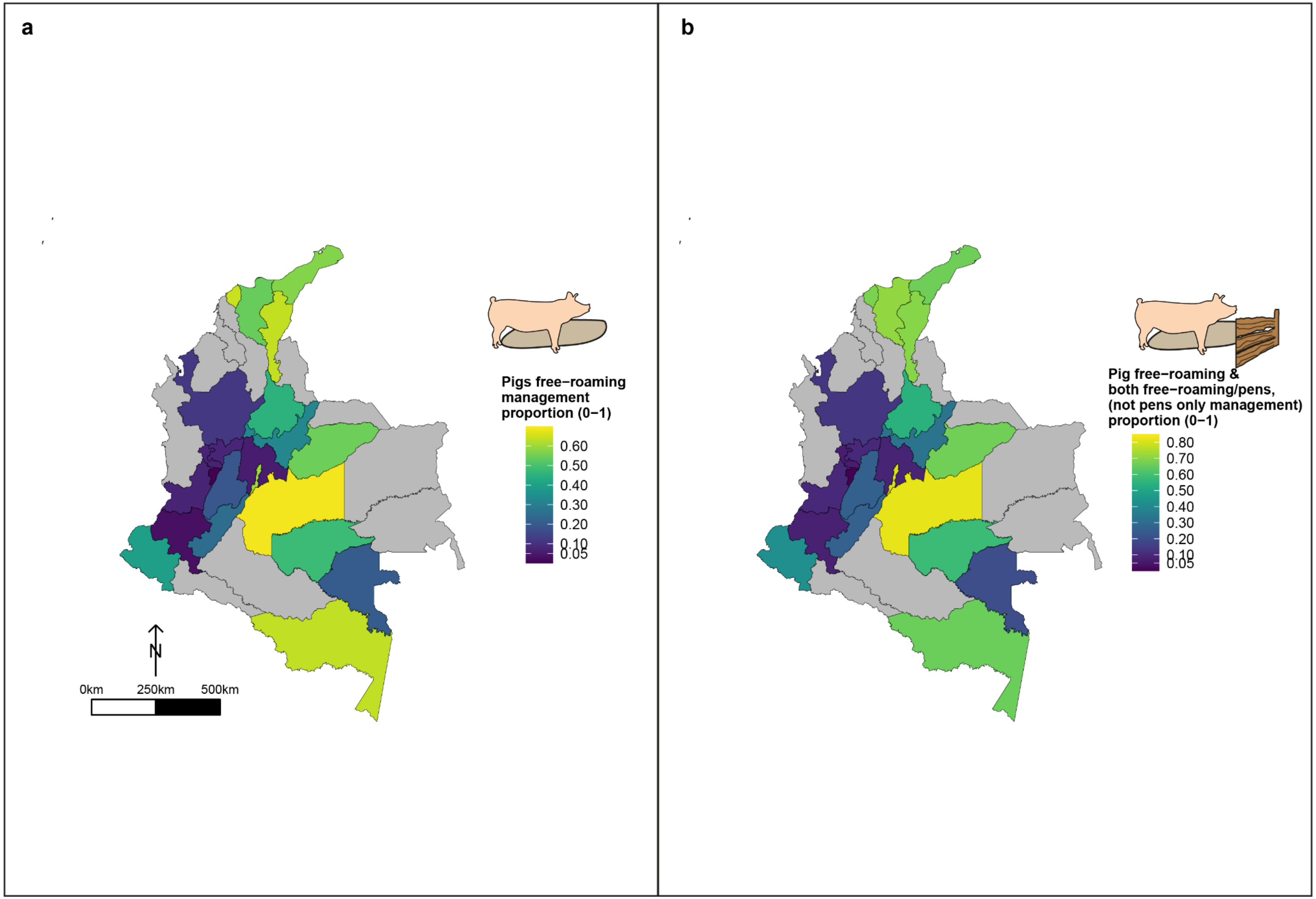


**Supplementary Figure S4.** Geographic variation in, (a) proportion of pigs which free-roam as a management strategy and (b) proportion of pigs which free roam or are held in mixed systems as a management strategy. These proportions are for a subset of the total dataset, only analysing those owning pigs (n=3,157)

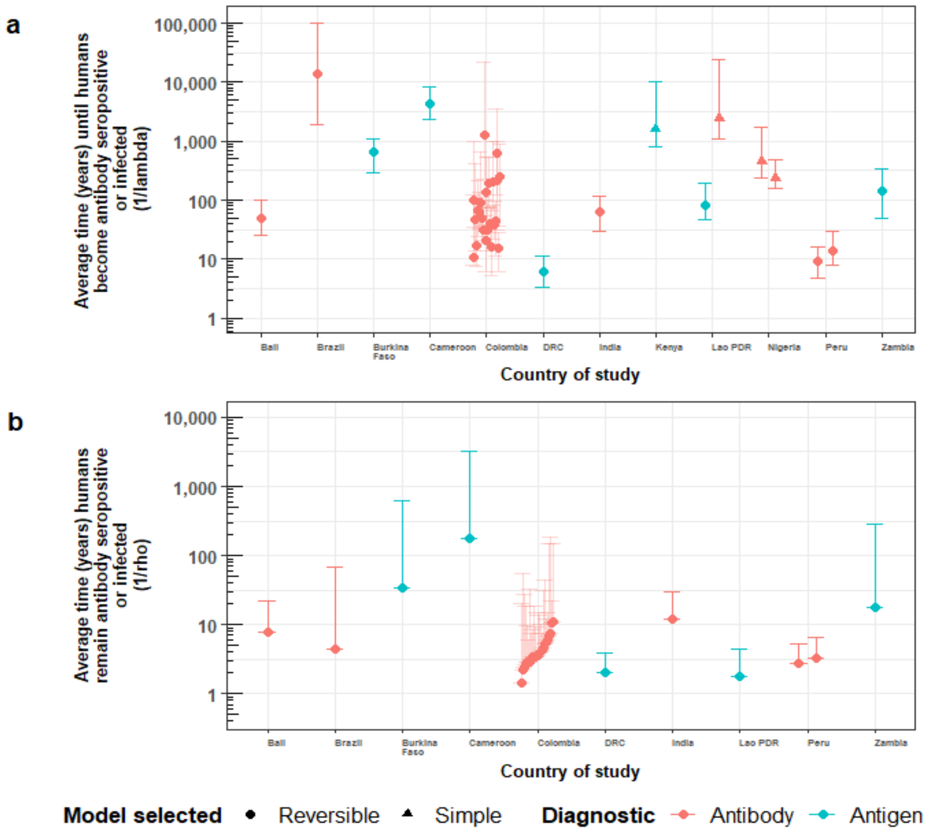


**Supplementary Figure S5. Country-specific estimates of (a) the average time (in years) until humans become cysticercosis antibody (Ab)-seropositive or infected ($1/\lambda$, vertical axis), and (b) the average time (in years) humans remain HCC Ab-seropositive or infected ($1/\rho$, vertical axis).** Marker colour denotes: red = Ab-seroprevalence; blue = antigen seroprevalence. Solid diamonds denote the use of the reversible catalytic model; circles are for the simple (Ab-seroconversion-only) model. Error bars are 95% Bayesian Credible Intervals around estimates. Colombia department error-bars are translucid to improve visibility of median estimates.

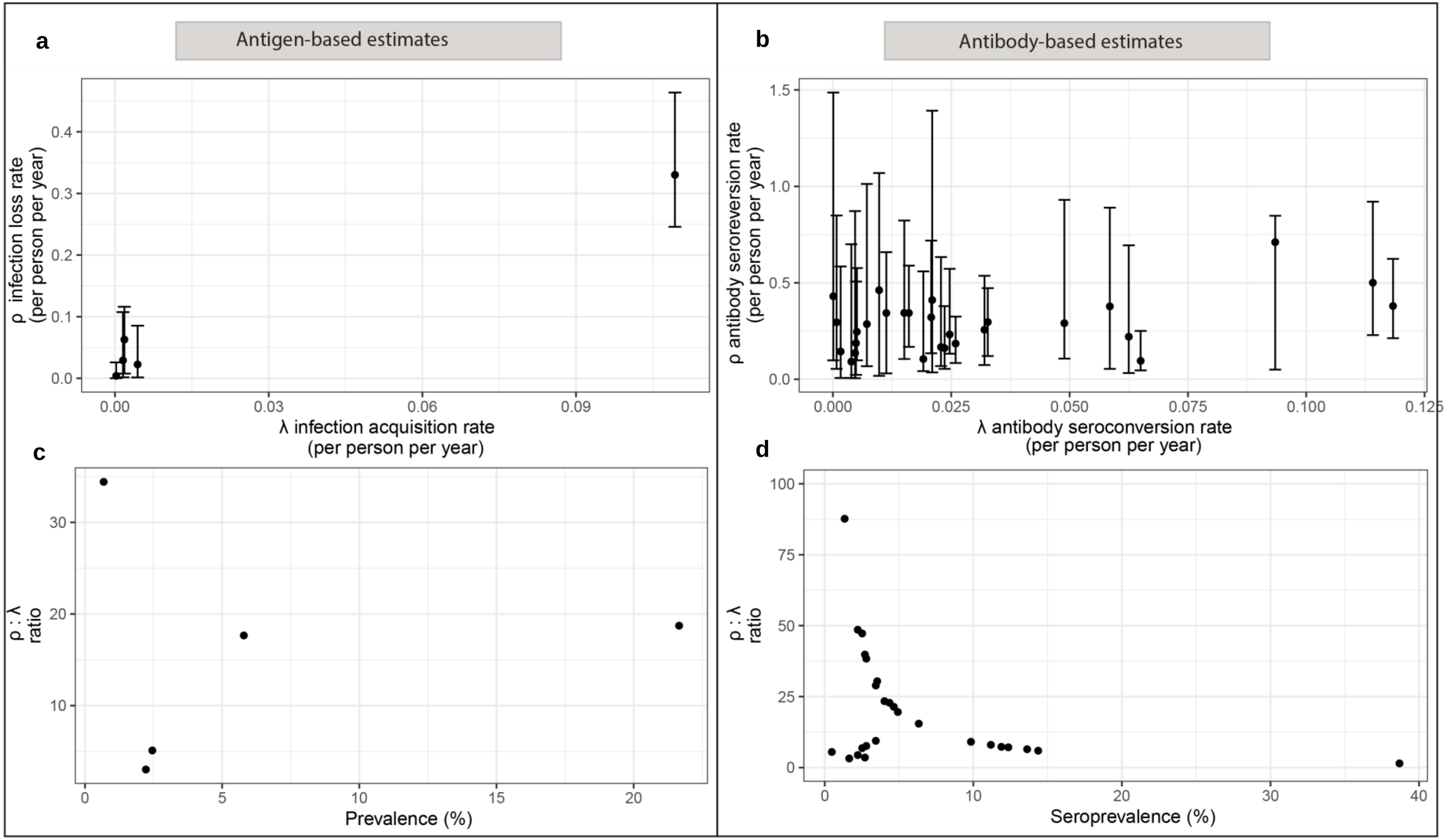


**Supplementary Figure S6.** Trends in *ρ* for increasing force-of-infection (FoI; *λ*) estimates for antigen-based estimates (a) and antibody (Ab)-based estimates (b), and trends in the infection loss ($\rho_{inf}$) to infection acquisition ($\lambda_{inf}$) rates by increasing prevalence for antigen-based estimates (c) and trends in Ab-seroreversion ($\rho_{sero}$) to Ab-seroconversion ($\lambda_{sero}$) rates by increasing Ab-seroprevalence for Ab-based estimates (d). The bottom panels show whether the magnitude in difference between *ρ: λ* with increasing (sero)prevalence (as a proxy for transmission intensity).

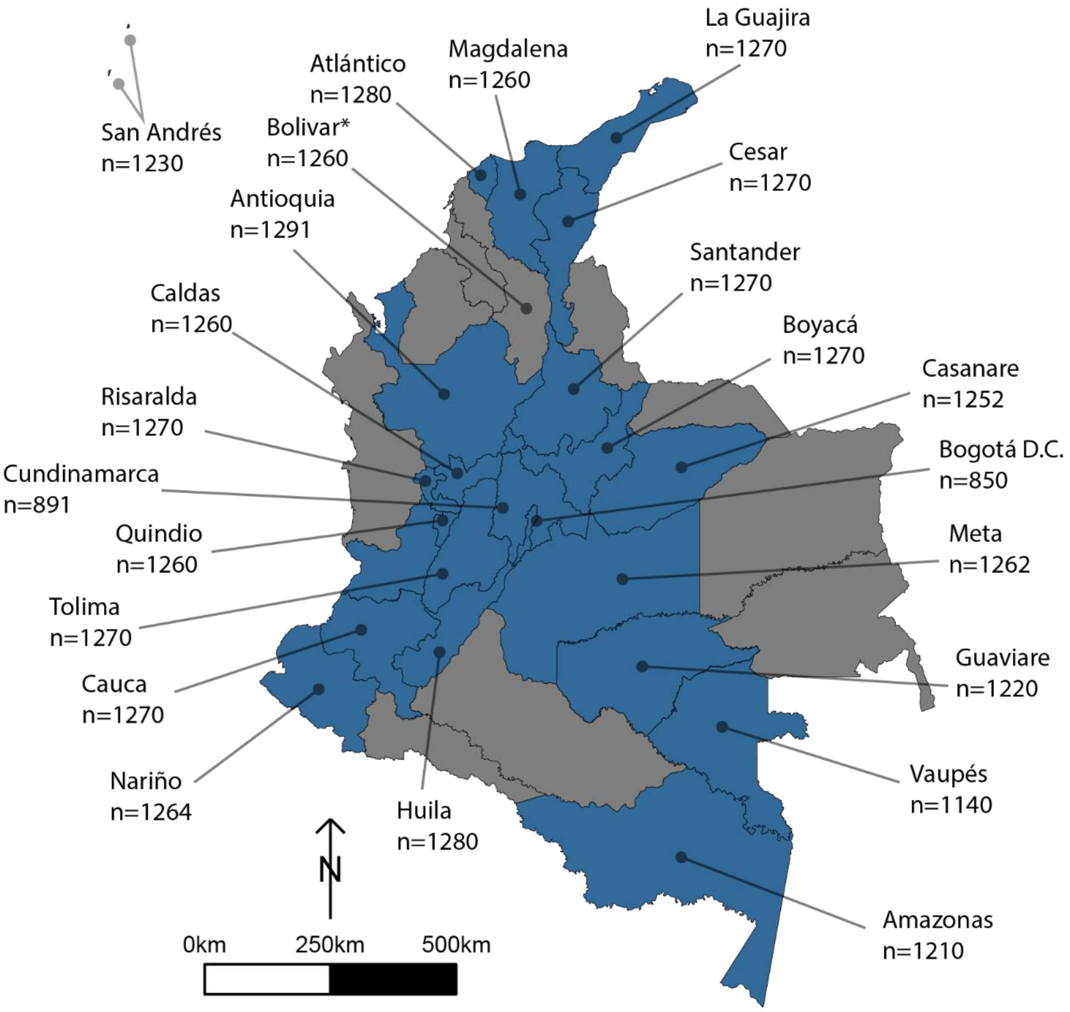


**Supplementary Figure S7. Human cysticercosis antibody serosurvey sample size by department in Colombia.** Blue departments are those included in the model fitting for this chapter. Bolivar* was not included in the model fitting (hence the Department is coloured in grey) - a full assessment is provided in the Results/Discussion of the main text regarding the exclusion of Bolivar. Data from Flórez Sánchez et al. (2013) and the Instituto National de Salud (INS).

a)

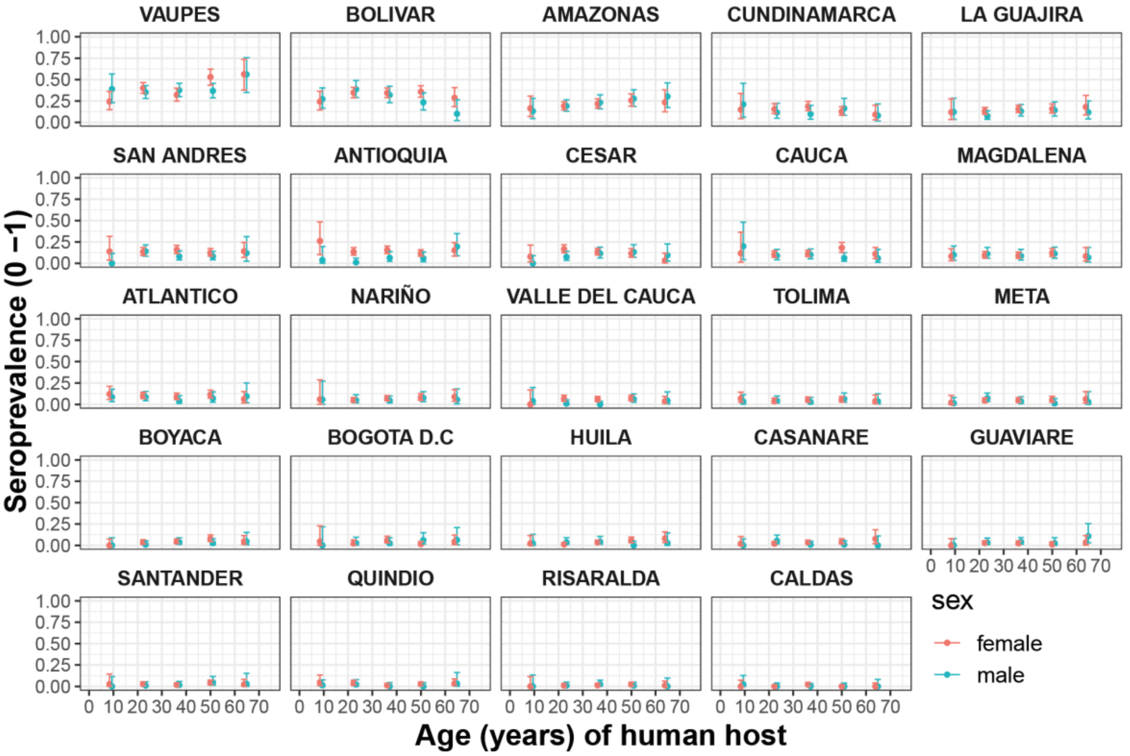


b)

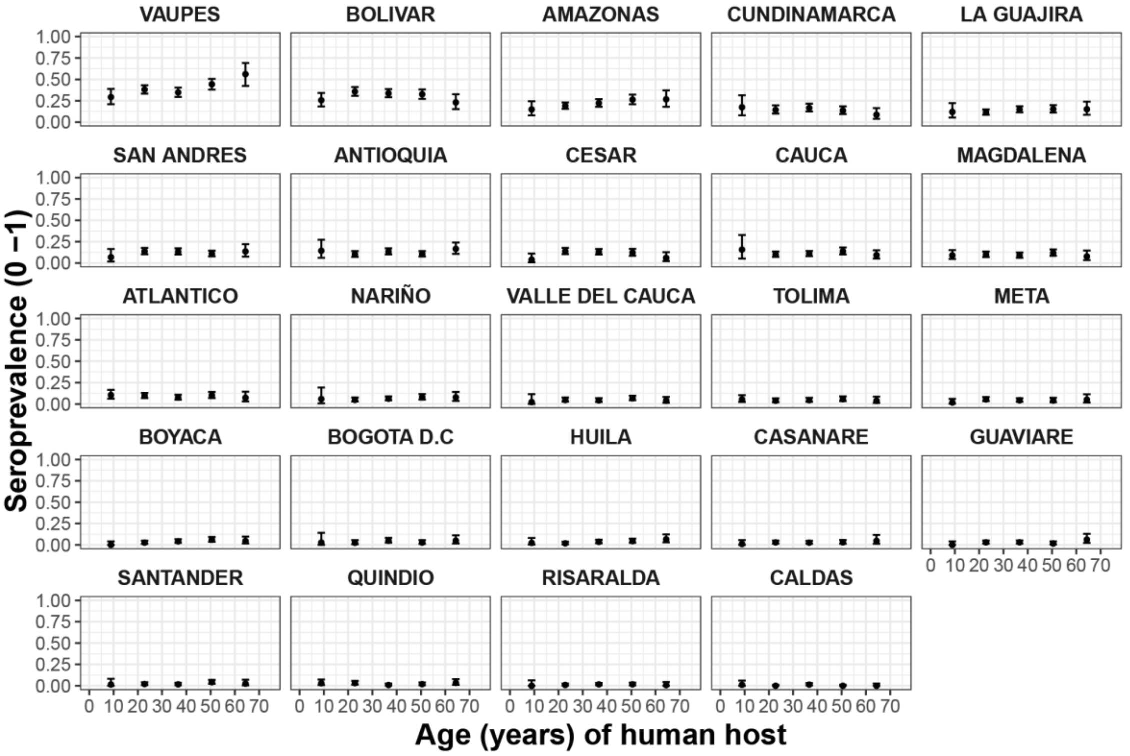


**Supplementary Figure S8.** Colombia (a) age- antibody (Ab) seroprevalence profile stratified by sex for each department (to determine whether profiles clearly differ by sex) and compared to (b) age- Ab-seroprevalence (non-sex stratified) profiles for all 24 departments.

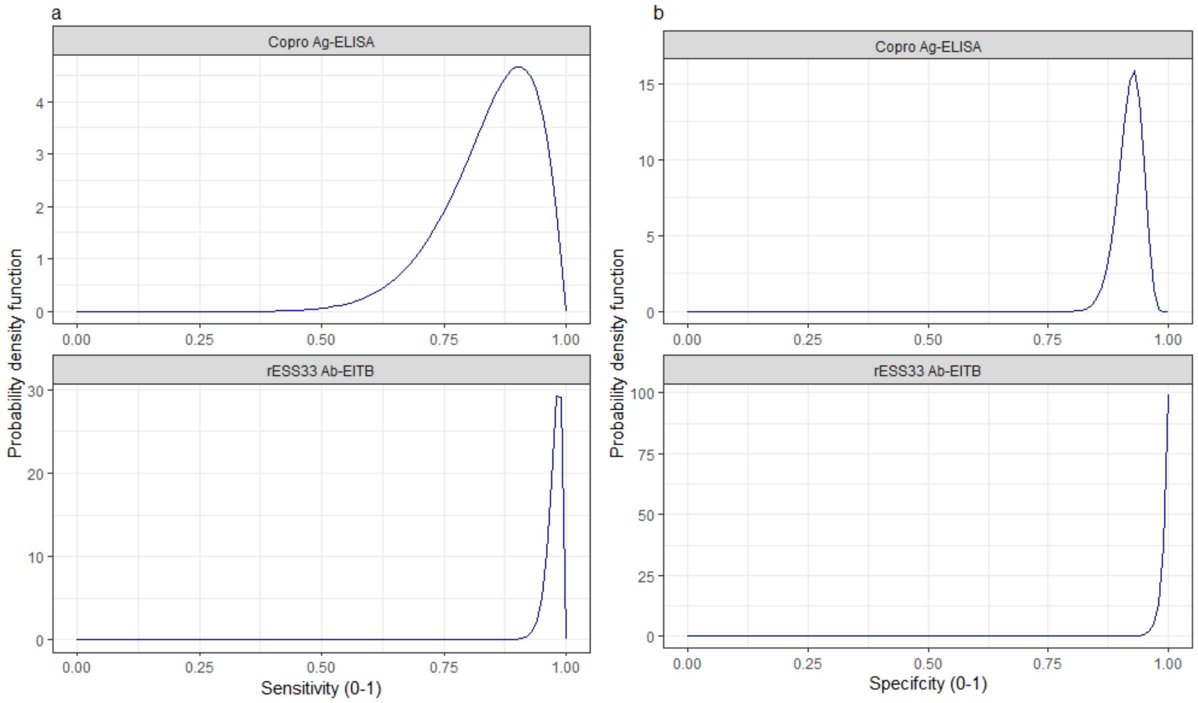


**Supplementary Figure S9. Informative beta distribution priors constructed for sensitivity and specificity parameters for each human taeniasis diagnostic**. $\beta$ prior distributions for: (a) sensitivity (se) (left-hand column plots); (b) specificity (sp) (right-hand column plots) of each diagnostic, constructed with $\alpha$ and $\beta$ shape parameters provided in Table 5.3. in Results (main text). The $\beta$ distribution provides a more flexible alternative to the uniform distribution, where the parameters of interest are constrained between 0 and 1. The shape parameters were chosen based on the literature estimates of se and sp (whereby $\alpha/(\alpha+\beta)$) equals the mean of the distribution (Lambert, 2018), and adapted where required to ensure that the spread of the prior distribution included the 95% confidence intervals identified in the literature. Ag: antigen; ELISA: enzyme-linked immunosorbent assay; Ab: antibody; EITB: enzyme-linked immunoelectrotransfer blot.

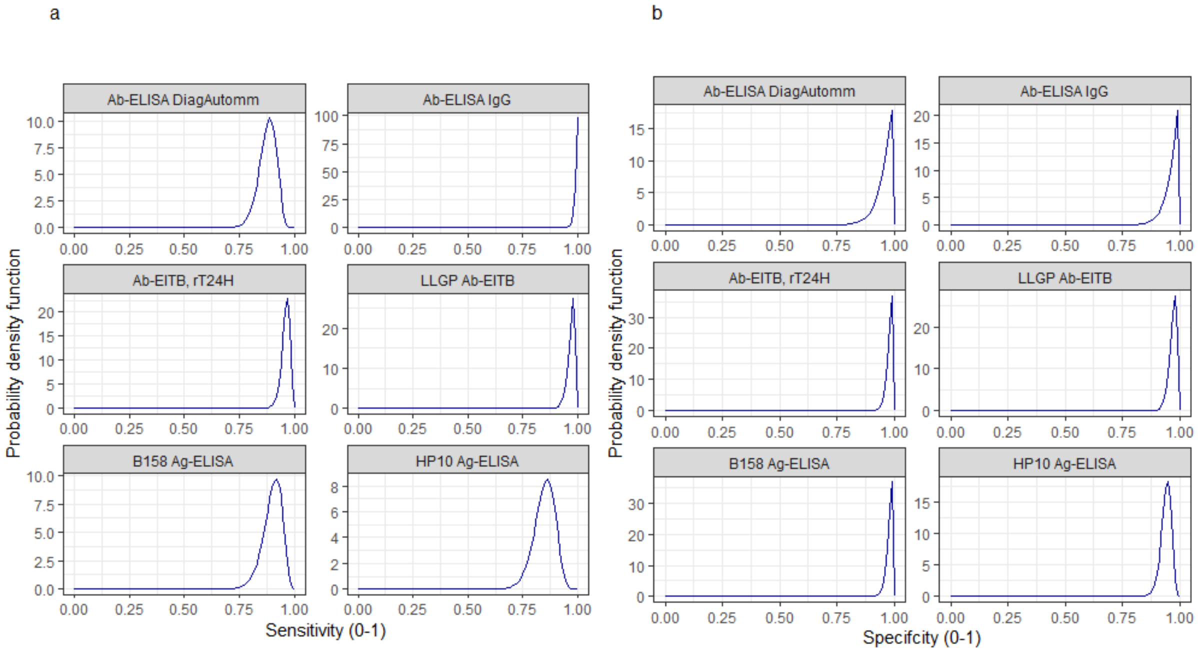


**Supplementary Figure S10. Informative beta distribution priors constructed for sensitivity and specificity parameters for each human cysticercosis diagnostic**. *β* prior distributions for: (a) sensitivity (se) and (b) specificity (sp) of each diagnostic, constructed with *α* and *β* shape parameters provided in Table 5.3. in Results (main text). The *β* distribution provides a more flexible alternative to the uniform distribution where the parameters of interest are constrained between 0 and 1 (Lambert, 2018). The shape parameters were chosen based on the literature estimates of se and sp (whereby $\alpha/(\alpha+\beta)$) equals the mean of the distribution (Lambert, 2018)), and adapted where required to ensure that the spread of the prior distribution included the 95% confidence intervals identified in the literature. Ab: antibody; ELISA: enzyme-linked immunosorbent assay; EITB: enzyme-linked immunoelectrotransfer blot; LLGP: lentil lectin-purified glycoprotein; Ig: immunoglobulin; Ag: antigen.

### Supplementary Tables

**Supplementary Table S1.** Summary of studies included in final analysis and the diagnostic parameters used to set the probabilistic constraints for sensitivity and specificity of each test.

| **Study author, year and supplementary reference** | **Location, country** | **Diagnostic** | **Sensitivity (%); specificity (%) median (95% confidence intervals given in the literature)** | **$\alpha$, $\beta$ shape parameters to construct each Beta distribution for sensitivity (Se) and specificity (Sp) priors (informed by column 4)** | **Total sample size** |
|---|---|---|---|---|---|
| Human taeniasis (copro-antigen and antibody) | | | | | |
| Gomes et al. (2002) | Mulungu do Morro, Brazil | Copro-antigen (Ag) enzyme-linked immunosorbent assay (ELISA) (Allan et al., 1990)* | 84.5 (61.9 – 98.0)<br>92.0 (90.0 – 93.8)[†] | Se: 11.5, 2.11<br>Sp: 100, 8.7 | 576 |
| Mwape et al. (2012) | Petauke district, Zambia | Copro-Ag ELISA (Allan et al., 1990) | 84.5 (61.9 – 98.0)<br>92.0 (90.0 – 93.8)[†] | Se: 11.5, 2.11<br>Sp: 100, 8.7 | 712 |
| Madinga et al. (2017) | Kimpese, DRC | Copro-Ag ELISA (Allan et al., 1990) | 84.5 (61.9 – 98.0)<br>92.0 (90.0 – 93.8)[†] | Se: 11.5, 2.11<br>Sp: 100, 8.7 | 4599 |
| Holt et al. (2016) [‡‡] | Luang Prabang and Savannakhet Provinces, Lao PDR | rES33-immunoblot (antibody) (Wilkins et al., 1999) | 97.6 (94.0 – 99.0)<br>99.0 (97.2 – 99.0) | Se: 100, 2.46<br>Sp: 99, 1 | 766 |
| **Human cysticercosis (antibody)** | | | | | |
| Theis et al. (1994) | Bali | Lentil lectin-purified glycoprotein enzyme-linked immunoelectrotransfer blot (LLGP-EITB) (Tsang et al., 1989) | 97.0 (95.0 – 1.00)<br>97.0 (94.0 – 99.0)[††] | Se: 100, 3.1<br>Sp: 100, 3.1 | 820 |

| | | | | | |
|---|---|---|---|---|---|
| Gomes et al. (2002) | Mulungu do Morro, Brazil | LLGP-EITB (Tsang et al., 1989) | 97.0 (95.0 – 1.00)<br>97.0 (94.0 – 99.0)[††] | Se: 100, 3.1<br>Sp: 100, 3.1 | 668 |
| Moro et al. (2003) | Vichaycocha, Peru | LLGP-EITB (Tsang et al., 1989) | 97.0 (95.0 – 1.00)<br>97.0 (94.0 – 99.0)[††] | Se: 100, 3.1<br>Sp: 100, 3.1 | 317 |
| Lescano et al. (2009) | Tumbes, Peru | LLGP-EITB (Tsang et al., 1989) | 97.0 (95.0 – 1.00)<br>97.0 (94.0 – 99.0)[††] | Se: 100, 3.1<br>Sp: 100, 3.1 | 738 |
| Jayaraman et al. (2011) | Vellore district, India | LLGP-EITB (Tsang et al., 1989) | 97.0 (95.0 – 1.00)<br>97.0 (94.0 – 99.0)[††] | Se: 100, 3.1<br>Sp: 100, 3.1 | 1056 |
| Weka et al. (2013) | Jos metropolis, Nigeria | IgG Ab-ELISA (DiagnosticAutomation/ CortezDiagnostic, Inc. (2016)) | 87.8 (79.0 – 93.9)<br>95.8 (86.0 – 99.0) | Se: 58.4, 8.1<br>Sp: 26.7, 1.2 | 125 |
| Edia-Asuke et al. (2015) | Kaduna metropolis, Nigeria | IgG Ab-ELISA (DiagnosticAutomation/ CortezDiagnostic, Inc. (2016)) | 87.8 (79.0 – 93.9)<br>95.8 (86.0 – 99.0) | Se: 58.4, 8.1<br>Sp: 26.7, 1.2 | 296 |
| Holt et al. (2016) [‡‡] | Luang Prabang and Savannakhet Provinces, Lao PDR | rT24H-immunoblot (antibody) (Hancock et al., 2006) | 96.0 (93.0 – 99.0)<br>98.0 (96.0 – 1) | Se: 100, 2.5<br>Sp: 100, 1 | 744 |
| Flórez Sánchez et al. (2013) | 24 departments across Colombia | IgG Ab-ELISA (López et al., 1988) | 1 (95% CI not provided)<br>97.6 (95% CI not provided) | Se: 98,1<br>Sp: 29.28, 1.08[‡] | 29360 |

| | | | | | |
|---|---|---|---|---|---|
| **Human cysticercosis (antigen)** | | | | | |
| Nguekam et al. (2003) | Menoua Division, Cameroon | B158/B60 Ag-ELISA; antigen ELISA using monoclonal antibodies vs excretory-secretory glycoproteins of *T. saginata* (Brandt et al., 1992; Dorny et al., 2000) | 90.0 (80.0 – 99.0)<br>98.0 (97.0 – 99.0)** | Se: 41.8, 4.6<br>Sp: 100, 2.04 | 4993 |
| Kanobana et al. (2011) | Malanga, DRC | B158/B60 Ag-ELISA (Brandt et al., 1992; Dorny et al., 2000) | 90.0 (80.0 – 99.0)<br>98.0 (97.0 – 99.0)** | Se: 41.8, 4.6<br>Sp: 100, 2.04 | 905 |
| Mwape et al. (2012) | Petauke district, Zambia | B158/B60 Ag-ELISA (Brandt et al., 1992; Dorny et al., 2000) | 90.0 (80.0 – 99.0)<br>98.0 (97.0 – 99.0)** | Se: 41.8, 4.6<br>Sp: 100, 2.04 | 708 |
| Conlan et al. (2012) | Four provinces, Lao PDR | B158/B60 Ag-ELISA (Brandt et al., 1992; Dorny et al., 2000) | 90.0 (80.0 – 99.0)<br>98.0 (97.0 – 99.0)** | Se: 41.8, 4.6<br>Sp: 100, 2.04 | 1306 |
| Sahlu et al. (2019) | 60 villages across three provinces in Burkina Faso | B158/B60 Ag-ELISA (Brandt et al., 1992; Dorny et al., 2000) | 90.0 (80.0 – 99.0)<br>98.0 (97.0 – 99.0)** | Se: 41.8, 4.6<br>Sp: 100, 2.04 | 2933 |
| Wardrop et al. (2015) [‡‡‡] | Lake Victoria crescent area, Kenya | HP10 Ag-ELISA; antigen ELISA using monoclonal antibodies vs excretory-secretory glycoproteins of *Taenia saginata* (Harrison et al., 1989) | 84.8 (74.4 – 95.2)<br>94 (90.2 – 97.8) [ǂ] | Se : 47.5, 8.5<br>Sp : 100, 6.4 | 2089 |

[†]sensitivity and specificity estimated in a Bayesian framework by Praet et al. (2013); [††] sensitivity and specificity based on exposure estimated in a Bayesian framework by Praet et al. (2010); *limited information on the specific protocol used for the copro-antigen assay; * sensitivity and specificity analysed by Noh et al. (2014); ** sensitivity and specificity estimated in a Bayesian framework by Praet et al. (2010); [‡] 95%CIs not provided to inform priors, therefore minimal uncertainty introduced around the sensitivity and specificity estimates to construct priors. Original data for two datasets available (under the Creative Commons Attribution License; CC BY 4.0) from the [‡‡]International Livestock Research Institute open-access repository (http://data.ilri.org/portal/dataset/ecozd) referenced in Holt et al. (2016) and [‡‡‡]University of Liverpool open-access repository (http://datacat.liverpool.ac.uk/352/) referenced in Fèvre et al. (2017); [ǂ] sensitivity and specificity estimated in Fleury et al. (2007).

DRC: Democratic Republic of the Congo; LPDR: Lao People's Democratic Republic; Ag; antigen; Ab: antibody; ELISA: enzyme-linked immunosorbent assay; LLGP-EITB: Lentil lectin-purified glycoprotein enzyme-linked immunoelectrotransfer blot; Ig: immunoglobulin.

**Supplementary Table S2. Observed prevalence estimates from sub-Saharan Africa, South America, and Asia for studies referring to "hyper-"or "highly-"endemic setting for human taeniasis (HTT) and human cysticercosis (HCC).**

| Reference | Location | Prevalence (%) | | | |
|---|---|---|---|---|---|
| | | HTT antibody | HTT Copro-Ag (coprology) | HCC antibody | HCC antigen |
| **sub-Saharan Africa** | | | | | |
| Mwanjali et al. (2013) | Tanzania | 4.1* | 5.2 (1.1) | 45.3** | 16.7[†] |
| Madinga et al. (2017) | DRC | | 23.4 | | |
| Mwape et al. (2012) | Zambia | | 6.3 | | 5.8[†] |
| **South America** | | | | | |
| Garcia et al. (2003) | Peru | | 2.8 | 13.1[††] | |
| Moyano et al. (2016) | Peru | | | 36.9[††] | |
| **Asia** | | | | | |
| Sato et al. (2018) | Lao PDR | 7.2[‡] | 3.1[‡‡] | | |
| Okello et al. (2014) | Lao PDR | | 26.1 | 66.7[††] | |

* rES38 Ab-immunblot (Wilkins et al., 1999); ** rT24H Ab-ELISA (Hancock et al., 2006); [†]B158/B60 Ag-ELISA (Brandt et al., 1992; Dorny et al., 2000); [††] LLGP-EITB (Tsang et al., 1989); [‡]Ab-ELISA confirmed by immunoblot (Sato et al., 2018); [‡‡]Copro-PCR (Sato et al., 2018).
DRC: Democratic Republic of the Congo (DRC); Lao PDR: Lao People's Democratic Republic.

**Supplementary Table S3. The deviance information criterion (DIC) and parameter estimates for simple and reversible catalytic models fitted to each observed human taeniasis (antibody and antigen) age-seroprevalence dataset (ordered by decreasing value of all-age seroprevalence).** For diagnostic methods used see the corresponding study in Supplementary Table S1.

| Dataset | All-age observed sero-prevalence (%) (95% CI) | Catalytic model | DIC value | Diagnostic sensitivity (95% BCI) | Diagnostic specificity (95% BCI) | $\lambda$ = infection acquisition ($\lambda_{inf}$) or seroconversion ($\lambda_{sero}$) rate, year$^{-1}$ (95% BCI) | 1/$\lambda$ = average time until becoming antibody seropositive (1/ $\lambda_{sero}$) or infected (1/ $\lambda_{inf}$) (years), (95% BCI) | $\rho$ = infection loss ($\rho_{inf}$) or seroreversion ($\rho_{sero}$) rate, year$^{-1}$ (95% BCI) | 1/$\rho$ = average time humans remain seropositive (1/ $\lambda_{sero}$) or infected (1/ $\lambda_{inf}$) (years) (95% BCI) |
|---|---|---|---|---|---|---|---|---|---|
| Jointly-fitted datasets – Simple catalytic model* | | | | | | | | | |
| Mwape et al. (2012) Antigen | 6.32 (4.65 – 8.37) | Simple | 52.51 | 0.835 (0.510 –0.976) | 0.951 (0.937 – 0.964) | 0.00085 (0.00009– 0.0023) | 1,170.41 (431.96 – 11,290.49) | NA | NA |
| Gomes et al. (2002) Antigen | 4.51 (2.97 – 6.54) | Simple | | | | 0.00022 (0.00002 – 0.001) | 4,508.79 (975.98 – 53,292.80) | NA | NA |
| Jointly-fitted datasets – Reversible catalytic model* | | | | | | | | | |
| Mwape et al. (2012) Antigen | 6.32 (4.65 – 8.37) | Reversible | 50.42$^{\dagger}$ | 0.824 (0.533 – 0.972) | 0.959 (0.941 – 0.976) | 0.021 (0.0038 – 0.062) | 47.53 (16.22 – 260.75) | 0.768 (0.362 – 0.991) | 1.303 (1.01 – 2.76) |
| Gomes et al. (2002) Antigen | 4.51 (2.97 – 6.54) | Reversible | | | | 0.0096 (0.00072 – 0.032) | 104.31 (31.33 – 1383.94) | 0.731 (0.379 – 0.978) | 1.37 (1.02 – 2.64) |

| | | | | | | | | | |
|---|---|---|---|---|---|---|---|---|---|
| Individually-fitted datasets | | | | | | | | | |
| Holt et al. (2016) Antibody | 2.49 (1.51 – 3.87) | Simple | 37.01[††] | 0.982 (0.959 – 0.996) | 0.992 (0.978 – 0.999) | 0.00044 (0.000103 – 0.00082) | 2264.05 (1,215.35 – 9,696.49) | NA | NA |
| Holt et al. (2016) Antibody | 2.49 (1.51 – 3.87) | Reversible | 37.87 | 0.978 (0.936 – 0.995) | 0.993 (0.977 – 0.999) | 0.013 (0.0018 – 0.0304) | 74.94 (32.91 – 556.49) | 0.761 (0.376 – 0.987) | 1.31 (1.01 – 2.66) |

Seroprevalence results are accompanied by 95% confidence intervals (95% CI) calculated by the Clopper-Pearson exact method. Parameter median posterior estimates are presented with 95% Bayesian credible intervals (95% BCI) and Deviance information criterion (DIC) model fitting scores;

* Diagnostic sensitivity and specificity jointly fitted for the Copro-Ag ELISA [8].

[†] Best-fitting model determined by DIC (jointly-fitted dataset). [††] Best-fitting model determined by DIC (individually-fitted dataset).

NA = Not applicable.

**Supplementary Table S4. The deviance information criterion (DIC) and parameter estimates for simple and reversible catalytic models fitted to each observed human cysticercosis antibody age-seroprevalence dataset (ordered by decreasing value of all-age seroprevalence).** For diagnostic methods used see the corresponding study in Supplementary Table S1.

| Dataset | All-age observed sero-prevalence (%) (95% CI) | Catalytic model | DIC value | Diagnostic sensitivity (95% BCI) | Diagnostic specificity (95% BCI) | $\lambda_{sero}$ = seroconver-sion rate, year$^{-1}$ (95% BCI) | $1/\lambda_{sero}$ = average time until becoming antibody seropositive (years) (95% BCI) | $\rho_{sero}$ = seroreversion rate, year$^{-1}$ (95% BCI) | $1/\rho_{sero}$ = average time humans remain antibody seropositive (years) (95% BCI) |
|---|---|---|---|---|---|---|---|---|---|
| Jointly fitted datasets – Simple catalytic model* | | | | | | | | | |
| Lescano et al. (2009) | 24.66 (21.59 – 27.94) | Simple | 259.32 | 0.969 (0.930 – 0.991) | 0.929 (0.911 – 0.946) | 0.00829 (0.0063 – 0.011) | 120.63 (90.91 – 158.73) | NA | NA |
| Moro et al. (2003) | 20.82 (16.48 – 25.71) | Simple | | | | 0.00311 (0.0015 – 0.0051) | 321.49 (197.84 – 663.30) | NA | NA |
| Jayaraman et al. (2011) | 15.81 (13.66 – 18.16) | Simple | | | | 0.0032 (0.0022 – 0.0043) | 312.00 (231.28 – 457.19) | NA | NA |
| Theis et al. (1994) | 12.68 (10.48 – 15.16) | Simple | | | | 0.00348 (0.0018 – 0.0054) | 287.22 (185.16 – 558.35) | NA | NA |
| Gomes et al. (2002) | 1.64 (0.82 – 2.93) | Simple | | | | 0.000078 (0.000012 – 0.00035) | 12,892.13 (2,854.80 – 82,689.12) | NA | NA |

| | | | | | | | | | |
|---|---|---|---|---|---|---|---|---|---|
| Jointly-fitted datasets – Reversible catalytic model* | | | | | | | | | |
| Lescano et al. (2009) | 24.66 (21.59 – 27.94) | Reversible | 156.28† | 0.976 (0.937 – 0.994) | 0.980 (0.967 – 0.988) | 0.12 (0.067 – 0.19) | 8.45 (5.20 – 15.03) | 0.38 (0.21 – 0.62) | 2.63 (1.60 – 4.69) |
| Moro et al. (2003) | 20.82 (16.48 – 25.71) | Reversible | | | | 0.11 (0.0504 – 0.21) | 8.77 (4.77 – 19.82) | 0.501 (0.23 – 0.92) | 2.00 (1.09 – 4.37) |
| Jayaraman et al. (2011) | 15.81 (13.66 – 18.16) | Reversible | | | | 0.019 (0.0095 – 0.093) | 52.32 (10.81 – 105.54) | 0.105 (0.042 – 0.56) | 9.51 (1.79 – 23.70) |
| Theis et al. (1994) | 12.68 (10.48 – 15.16) | Reversible | | | | 0.024 (0.011 – 0.052) | 42.38 (19.47 – 92.51) | 0.16 (0.054 – 0.38) | 6.19 (2.64 – 18.57) |
| Gomes et al. (2002) | 1.64 (0.82 – 2.93) | Reversible | | | | 0.000086 (0.000011 – 0.00066) | 11,595.71 (1,519.803 – 91,577.104) | 0.43 (0.098 – 1.49) | 2.32 (0.67 – 10.19) |
| Jointly fitted datasets – Simple catalytic model** | | | | | | | | | |
| Edia-Asuke et al. (2015) | 14.53 (10.72 – 19.06) | Simple | 38.21† | 0.872 (0.784 – 0.942) | 0.974 (0.916 – 0.998) | 0.0044 (0.0018 – 0.0064) | 226.98 (156.27 – 507.04) | NA | NA |
| Weka et al. (2013) | 9.60 (5.06 – 16.17) | Simple | | | | 0.0023 (0.00053 – 0.0046) | 434.95 (217.65 – 1,888.93) | NA | NA |

| | | | | | | | | | |
|---|---|---|---|---|---|---|---|---|---|
| Jointly fitted datasets – Reversible catalytic model** | | | | | | | | | |
| Edia-Asuke et al. (2015) | 14.53 (10.72 – 19.06) | Reversible | 54.62 | 0.880 (0.791 – 0.943) | 0.878 (0.846 – 0.919) | 0.0063 (0.00074 – 0.044) | 158.30 (22.54 – 1,356.21) | 5.55 (0.092 – 11.81) | 0.18 (0.08 – 10.88) |
| Weka et al. (2013) | 9.60 (5.06 – 16.17) | Reversible | | | | 0.0027 (0.00033 – 0.019) | 371.93 (53.69 – 3,050.51) | 6.37 (0.86 – 11.27) | 0.16 (0.09 – 1.16) |
| Individually-fitted datasets | | | | | | | | | |
| Holt et al. (2016) | 2.96 (1.86 – 4.44) | Simple | 36.62[††] | 0.964 (0.914 – 0.988) | 0.986 (0.969 – 0.997) | 0.00044 (0.000049 – 0.00090) | 2,289.04 (1,111.72 – 20,402.51) | NA | NA |
| Holt et al. (2016) | 2.96 (1.86 – 4.44) | Reversible | 48.34 | 0.963 (0.916 – 0.988) | 0.973 (0.960 – 0.984) | 0.00048 (0.000066 – 0.0038) | 2,077.84 (266.53 – 15,092.40) | 1.27 (0.099 – 2.55) | 0.79 (0.39 – 10.02) |

Seroprevalence results are accompanied by 95% confidence intervals (95% CI) calculated by the Clopper-Pearson exact method. Parameter median posterior estimates are presented with 95% Bayesian credible intervals (95% BCI) and Deviance information criterion (DIC) model fitting scores;
* Diagnostic sensitivity and specificity for the antibody lentil lectin-purified glycoprotein enzyme-linked immunoelectrotransfer blot (Ab LLGP-EITB) assay (Tsang et al., 1989) were jointly fitted across datasets.
** Diagnostic sensitivity and specificity for the antibody Ab-ELISA (DiagnosticAutomation/CortezDiagnostic (2006)) assay were jointly fitted across datasets.
[†] Best-fitting model determined by DIC (jointly-fitted dataset). [††] Best-fitting model determined by DIC (individually-fitted dataset).
NA = Not applicable.

**Supplementary Table S5. The deviance information criterion (DIC) and parameter estimates for simple and reversible catalytic models fitted to each observed human cysticercosis antigen age-seroprevalence dataset (ordered by decreasing value of all-age seroprevalence).** For diagnostic methods used see the corresponding study in Supplementary Table S1.

| **Dataset** | **All-age observed sero-prevalence (%) (95% CI)** | **Catalytic model** | **DIC value** | **Diagnostic sensitivity (95% BCI)** | **Diagnostic specificity (95% BCI)** | **$\lambda_{inf}$ = infection acquisition rate, year$^{-1}$ (95% BCI)** | **$1/\lambda_{inf}$ = average time until becoming antigen seropositive (years) (95% BCI)** | **$\rho_{inf}$ = infection loss rate, year$^{-1}$ (95% BCI)** | **$1/\rho_{inf}$ = average time humans remain antigen seropositive (years) (95% BCI)** |
|---|---|---|---|---|---|---|---|---|---|

Jointly fitted datasets – Simple catalytic model*

| **Dataset** | **All-age observed sero-prevalence (%) (95% CI)** | **Catalytic model** | **DIC value** | **Diagnostic sensitivity (95% BCI)** | **Diagnostic specificity (95% BCI)** | **$\lambda_{inf}$ = infection acquisition rate, year$^{-1}$ (95% BCI)** | **$1/\lambda_{inf}$ = average time until becoming antigen seropositive (years) (95% BCI)** | **$\rho_{inf}$ = infection loss rate, year$^{-1}$ (95% BCI)** | **$1/\rho_{inf}$ = average time humans remain antigen seropositive (years) (95% BCI)** |
|---|---|---|---|---|---|---|---|---|---|
| Kanobana et al. (2011) | 21.66 (19.01 – 24.49) | Simple | 309.87 | 0.843 (0.768 – 0.916) | 0.996 (0.992 – 0.999) | 0.011 (0.0088 – 0.013) | 92.29 (74.45 – 113.63) | NA | NA |
| Mwape et al. (2012) | 5.79 (4.19 – 7.77) | Simple | | | | 0.0031 (0.00203 – 0.0045) | 318.56 (222.77 – 463.60) | NA | NA |
| Sahlu et al. (2019) | 2.45 (1.93 – 3.08) | Simple | | | | 0.00099 (0.00071 – 0.0014) | 1,008.41 (732.52 – 1,409.59) | NA | NA |
| Conlan et al. (2012) | 2.22 (1.49 – 3.17) | Simple | | | | 0.000702 (0.0004 – 0.0011) | 1,424.92 (903.91 – 2,585.53) | NA | NA |
| Nguekam et al. (2003) | 0.68 (0.47 – 0.95) | Simple | | | | 0.00017 (0.000078 – 0.00029) | 5,720.002 (3,376.93 – 12,874.48) | NA | NA |

| | | | | | | | | | |
|---|---|---|---|---|---|---|---|---|---|
| Jointly fitted datasets – Reversible catalytic model* | | | | | | | | | |
| Kanobana et al. (2011) | 21.66 (19.01 – 24.49) | Reversible | 227.06† | 0.909 (0.810 – 0.967) | 0.999 (0.995 – 0.999) | 0.11 (0.077 – 0.17) | 9.15 (5.91 – 13.03) | 0.330 (0.246 – 0.464) | 3.03 (2.16 – 4.07) |
| Mwape et al. (2012) | 5.79 (4.19 – 7.77) | Reversible | | | | 0.0044 (0.0027 – 0.0088) | 226.55 (113.85 –375.75) | 0.023 (0.001 – 0.085) | 44.42 (11.72 – 672.09) |
| Sahlu et al. (2019) | 2.45 (1.93 – 3.08) | Reversible | | | | 0.0016 (0.00091 – 0.0032) | 642.11 (308.19 – 1,106.82) | 0.029 (0.0016 – 0.11) | 34.31 (9.31 – 624.45) |
| Conlan et al. (2012) | 2.22 (1.49 – 3.17) | Reversible | | | | 0.0018 (0.00073 – 0.0034) | 547.64 (298.86 – 1,371.48) | 0.063 (0.0076 – 0.12) | 15.91 (8.62 – 130.78) |
| Nguekam et al. (2003) | 0.68 (0.47 – 0.95) | Reversible | | | | 0.00017 (0.000077 – 0.00030) | 4,376.16 (2,617.01 – 7,679.21) | 0.004 (0.00018 – 0.026) | 247.89 (38.62 – 5,600.50) |
| Individually-fitted datasets | | | | | | | | | |
| Wardrop et al. (2015) | 6.61 (5.57 – 7.76) | Simple | 79.55†† | 0.850 (0.735 – 0.927) | 0.944 (0.930 – 0.959) | 0.00054 (0.000053 – 0.0013) | 1,868.83 (803.17 – 18,740.50) | NA | NA |
| Wardrop et al. (2015) | 6.61 (5.57 – 7.76) | Reversible | 92.25 | 0.854 (0.752 – 0.928) | 0.935 (0.924 – 0.946) | 0.00059 (0.000079 – 0.0045) | 1,692.74 (220.57 – 12,631.65) | 1.68 (0.12 – 4.31) | 0.596 (0.23 – 8.07) |

Seroprevalence results are accompanied by 95% confidence intervals (95% CI) calculated by the Clopper-Pearson exact method. Parameter median posterior estimates are presented with 95% Bayesian credible intervals (95% BCI) and Deviance information criterion (DIC) model fitting scores;
* Diagnostic sensitivity and specificity for the B158/B60 Ag-ELISA (Brandt et al., 1992; Dorny et al., 2000).
† Best-fitting model determined by DIC (jointly-fitted dataset). †† Best-fitting model determined by DIC (individually-fitted dataset).
NA = Not applicable.

**Supplementary Table S6. The deviance information criterion (DIC) and parameter estimates for the reversible catalytic model jointly fitted (for diagnostic sensitivity and specificity) to the observed human cysticercosis antibody age-seroprevalence for each available department in Colombia (n=23, ordered by decreasing value of all-age seroprevalence).** For diagnostic methods used see the corresponding study in Supplementary Table S1.

| **Department (sample size, n)** | **All-age observed seroprevalence (%) (95% CI)** | **$\lambda_{sero}$ = seroconver-sion rate, $year^{-1}$ (95% BCI)** | **$1/\lambda_{sero}$ = average time until becoming antibody seropositive (years) (95% BCI)** | **$\rho_{sero}$ = seroreversion rate, $year^{-1}$ (95% BCI)** | **$1/\rho_{sero}$ = average time humans remain antibody seropositive (years) (95% BCI)** |
|---|---|---|---|---|---|
| Vaupés (1,140) | 38.68 (35.85 – 41.58) | 0.065 (0.036 – 0.16) | 15.38 (6.24 – 27.53) | 0.095 (0.046 – 0.25) | 10.47 (3.99 – 21.77) |
| Amazonas (1,210) | 21.74 (19.44 – 24.17) | 0.063 (0.013 – 0.19) | 15.60 (5.24 – 74.20) | 0.22 (0.032 – 0.69) | 4.53 (1.44 – 30.78) |
| Cundinamarca (891) | 14.37 (12.13 – 16.84) | 0.049 (0.018 – 0.16) | 20.45 (6.06 – 55.00) | 0.29 (0.11 – 0.93) | 3.44 (1.08 – 9.34) |
| La Guajira (1,270) | 13.62 (11.78 – 15.63) | 0.059 (0.0097 – 0.13) | 17.09 (7.43 – 102.33) | 0.38 (0.054 – 0.89) | 2.64 (1.12 – 18.60) |
| San Andrés (1,230) | 12.36 (10.57 – 14.33) | 0.026 (0.012 – 0.047) | 38.56 (21.32 – 83.26) | 0.19 (0.084 – 0.32) | 5.40 (3.08 – 11.87) |
| Antioquia (1,291) | 12.01 (10.28 – 13.90) | 0.093 (0.0082 – 0.13) | 10.70 (7.80 – 121.99) | 0.71 (0.050 – 0.85) | 1.41 (1.18 – 19.91) |
| Cesar (1,270) | 11.89 (10.16 – 13.80) | 0.023 (0.010 – 0.087) | 43.77 (11.54 – 96.23) | 0.17 (0.068 – 0.63) | 5.99 (1.58 – 14.60) |
| Cauca (1,270) | 11.18 (9.50 – 13.04) | 0.032 (0.010 – 0.071) | 31.23 (14.13 – 97.66) | 0.26 (0.074 – 0.54) | 3.90 (1.86 – 13.55) |
| Magdalena (1,260) | 9.84 (8.25 – 11.62) | 0.033 (0.013 – 0.054) | 30.56 (18.48 – 74.19) | 0.30 (0.12 – 0.47) | 3.37 (2.12 – 8.31) |
| Atlántico (1,280) | 9.06 (7.55 – 10.77) | 0.025 (0.013 – 0.57) | 40.54 (17.47 – 77.42) | 0.23 (0.13 – 0.57) | 4.30 (1.75 – 7.54) |
| Nariño (1,264) | 6.33 (5.05 – 7.82) | 0.021 (0.0088 – 0.048) | 48.14 (20.87 – 113.25) | 0.32 (0.13 – 0.72) | 3.11 (1.39 – 7.42) |
| Valle Del Cauca (1,260) | 4.92 (3.79 – 6.26) | 0.021 (0.0024 – 0.072) | 47.67 (13.92 – 408.47) | 0.41 (0.036 – 1.39) | 2.44 (0.72 – 27.91) |
| Tolima (1,270) | 4.65 (3.56 – 5.95) | 0.016 (0.0074 – 0.031) | 62.27 (32.59 – 134.42) | 0.34 (0.17 – 0.59) | 2.91 (1.70 – 5.95) |
| Meta (1,262) | 4.36 (3.30 – 5.64) | 0.015 (0.0047 – 0.041) | 66.32 (24.27 – 212.47) | 0.34 (0.11 – 0.82) | 2.91 (1.22 – 9.51) |
| Boyacá (1,270) | 4.02 (3.00 – 5.25) | 0.0039 (0.0011 – 0.028) | 256.18 (35.80 – 872.09) | 0.091 (0.0068 – 0.70) | 10.95 (1.43 – 146.97) |
| Bogotá D.C (850) | 3.53 (2.39 – 5.00) | 0.011 (0.0015 – 0.027) | 88.55 (36.77 – 665.86) | 0.34 (0.030 – 0.66) | 2.91 (1.52 – 32.81) |
| Huila (1,280) | 3.43 (2.51 – 4.59) | 0.0047 (0.0010 – 0.032) | 212.91 (31.19 – 977.68) | 0.14 (0.0054 – 0.87) | 7.36 (1.15 – 183.64) |

| | | | | | |
|---|---|---|---|---|---|
| Casanare (1,252) | 2.80 (1.95 – 3.87) | 0.0049 (0.0010 – 0.015) | 204.76 (65.61 – 966.12) | 0.19 (0.023 – 0.51) | 5.34 (1.97 – 43.56) |
| Guaviare (1,220) | 2.70 (1.87 – 3.78) | 0.0072 (0.0019 – 0.026) | 139.16 (38.01 – 539.23) | 0.27 (0.068 – 1.01) | 3.49 (0.99 – 14.73) |
| Santander (1,270) | 2.52 (1.73 – 3.54) | 0.0098 (0.0010 – 0.029) | 102.34 (34.60 – 996.14) | 0.46 (0.018 – 1.070) | 2.17 (0.94 – 55.69) |
| Quindio (1,260) | 2.22 (1.48 – 3.20) | 0.0051 (0.0019 – 0.014) | 197.21 (72.80 – 537.87) | 0.25 (0.098 – 0.58) | 4.06 (1.74 – 10.20) |
| Risaralda (1,270) | 1.39 (0.78 – 2.13) | 0.0016 (0.00029 – 0.0081) | 609.19 (123.15 – 3,425.94) | 0.14 (0.0067 – 0.58) | 6.95 (1.71 – 148.42) |
| Caldas (1,260) | 0.48 (0.17-1.03) | 0.00079 (0.000045 – 0.0045) | 1,263.06 (224.61 – 22,321.43) | 0.30 (0.054 – 0.85) | 3.39 (1.18 – 18.47) |

**DIC score for the reversible model was -401.78. Jointly-fitted diagnostic sensitivity was 0.989 (95%BCI: 0.946 – 0.999) and specificity was 0.998 (95%BCI: 0.993 – 0.999).**
Seroprevalence results are accompanied by 95% confidence intervals (95% CI) calculated by the Clopper-Pearson exact method. Parameter median posterior estimates are presented with 95% Bayesian credible intervals (95% BCI) and Deviance information criterion (DIC) model fitting scores.

**Supplementary Table S7. The deviance information criterion (DIC) and parameter estimates for the simple catalytic model jointly fitted (for diagnostic sensitivity and specificity) to the observed human cysticercosis antibody age-seroprevalence for each available department in Colombia (n = 23, ordered by decreasing value of all-age seroprevalence**). For diagnostic methods used see the corresponding study in Supplementary Table S1.

| **Department (sample size, n)** | **All-age observed seroprevalence (%) (95% CI)** | **$\lambda_{sero}$ = seroconversion rate, year$^{-1}$ (95% BCI)** | **1/$\lambda_{sero}$ = average time until becoming antibody seropositive (years) (95% BCI)** |
|---|---|---|---|
| Vaupés (1,140) | 38.68 (35.85 – 41.58) | 0.014 (0.013 – 0.016) | 69.49 (62.75 – 77.02) |
| Amazonas (1210) | 21.74 (19.44 –24.17) | 0.0066 (0.0057 – 0.0075) | 151.98 (133.29 – 174.86) |
| Cundinamarca (891) | 14.37 (12.13 – 16.84) | 0.0034 (0.0027 – 0.0042) | 292.03 (246.92 – 366.08) |
| La Guajira (1270) | 13.62 (11.78 – 15.63) | 0.0036 (0.00029 – 0.0042) | 281.41 (236.63 – 340.74) |
| San Andrés (1230) | 12.36 (10.57 – 14.33) | 0.0029 (0.0024 – 0.0035) | 343.07 (283.07 – 419.36) |
| Antioquia (1291) | 12.01 (10.28 –13.90) | 0.0027 (0.0022 – 0.0033) | 366.35 (306.56 – 453.58) |
| Cesar (1270) | 11.89 (10.16 – 13.80) | 0.0029 (0.0026 – 0.0035) | 342.57 (282.18 – 424.10) |
| Cauca (1270) | 11.18 (9.50 – 13.04) | 0.0025 (0.0020 – 0.0031) | 399.19 (319.87 – 494.95) |
| Magdalena (1260) | 9.84 (8.25 – 11.62) | 0.0023 (0.0018 – 0.0028) | 439.17 (353.09 – 565.11) |
| Atlántico (1280) | 9.06 (7.55 – 10.77) | 0.0021 (0.0015 – 0.0026) | 485.98 (379.09 – 651.88) |
| Nariño (1264) | 6.33 (5.05 – 7.82) | 0.0012 (0.00080 – 0.0016) | 834.70 (1242.79 – 613.61) |
| Valle Del Cauca (1260) | 4.92 (3.79 – 6.26) | 0.00075 (0.00043 – 0.0011) | 1,327.11 (884.23 – 2,326.84) |
| Tolima (1270) | 4.65 (3.56 – 5.95) | 0.00070 (0.00037 – 0.0011) | 1,437.40 (918.07 – 2,736.72) |
| Meta (1262) | 4.36 (3.30 – 5.64) | 0.00067 (0.00036 – 0.0011) | 1,485.96 (919.29 – 2,825.13) |
| Boyacá (1270) | 4.02 (3.00 – 5.25) | 0.00060 (0.00029 – 0.00095) | 1,670.93 (1,052.29 – 3,441.97) |
| Bogotá D.C (850) | 3.53 (2.39 – 5.00) | 0.00041 (0.00013 – 0.00080) | 2,461.14 (1,250.71 – 7,784.22) |
| Huila (1280) | 3.43 (2.51 – 4.59) | 0.00046 (0.00018 – 0.00079) | 2,174.64 (1,261.06 – 5,466.00) |

| | | | |
|---|---|---|---|
| Casanare (1252) | 2.80 (1.95 – 3.87) | 0.00026 (0.000054 – 0.00057) | 3,786.45 (1,766.88 – 91,388.13) |
| Guaviare (1220) | 2.70 (1.87 – 3.78) | 0.00025 (0.000043 – 0.00055) | 3,994.80 (1,807.03 – 23,180.19) |
| Santander (1270) | 2.52 (1.73 – 3.54) | 0.00018 (0.000033 – 0.00046) | 5,065.60 (2,196.77 – 29,890.66) |
| Quindio (1260) | 2.22 (1.48 – 3.20) | 0.00011 (0.000015 – 0.00033) | 9,210.03 (3,062.74 – 66,454.01) |
| Risaralda (1270) | 1.39 (0.78 – 2.13) | 0.000055 (0.000012 – 0.00019) | 1,8304.61 (5,200.37 – 85,265.30) |
| Caldas (1260) | 0.48 (0.17 – 1.03) | 0.000031 (0.000011 – 0.00011) | 32,772.10 (8,874.67 – 91,388.13) |

**DIC score for the simple model was –442.7. Jointly-fitted diagnostic sensitivity was 0.987 (95%BCI: 0.966 – 0.999) and specificity was 0.980 (95%BCI: 0.975 – 0.984).**

Seroprevalence results are accompanied by 95% confidence intervals (95% CI) calculated by the Clopper-Pearson exact method. Parameter median posterior estimates are presented with 95% Bayesian credible intervals (95% BCI) and Deviance information criterion (DIC) model fitting scores.

**PRIME-NTD (Policy-Relevant Items for Reporting Models in Epidemiology of Neglected Tropical Diseases) Summary (Supplementary Table S8)**

| Principle | What has been done to satisfy the principle? | Where in the manuscript is this described? |
|---|---|---|
| **1. Stakeholder engagement** | Engagement with the *Instituto Nacional de Salud* in Colombia for access to a large dataset, and continued engagement with the stakeholders throughout the analysis & manuscript development. | Intro (pg.5); Discussion (pg.24-25). |
| **2. Complete model documentation** | Full model description in the manuscript and all model/ fitting code available on the Github link: *https://github.com/mrc-ide/human_tsol_FoI_modelling* | Methods (pg. 27-29) and "code availability" statement. |
| **3. Complete description of data used** | In-depth description of datasets and age-(sero)prevalence aggregated data available. | Methods (pg. 27), Supplementary material tables (pg. 13-15), and "data availability statement". |
| **4. Communicating uncertainty** | Methods for generating uncertainty associated with fitted model parameters are described, and Bayesian credible intervals presented in the results. Uncertainty is also interpreted in the Discussion. | Methods (pg. 28-29; including specifying prior distributions to sample uncertainty in Table 5.), throughout Results section, Discussion (pg. 23-24) |
| **5. Testable model outcomes** | No formal model validation exercises performed; however fitted parameter estimates discussed in terms of literature estimates. | Discussion (pg. 23) |

Full formulation of the principles:

1. Don't do it alone. Engage stakeholders throughout, from the formulation of questions to the discussions on the implications of the findings.
2. Reproducibility is key! Prepare and make available (preferably open-source) a complete technical documentation of all model code, mathematical formulas, assumptions and their justification, allowing others to reproduce the model.
3. Model calibration, goodness-of-fit and validation are fundamental processes of scientific modelling. All data used should be described in sufficient detail to allow the reader to assess the type and quality of these analyses. When using data by reference, use Principle 2.
4. Communicating uncertainty is a hallmark of good modelling practice. Perform a sensitivity analysis of all key parameters, and for each paper reporting model predictions include an uncertainty assessment of those model outputs within the paper.
5. Model outcomes should be articulated in the form of testable hypotheses. This allows comparison with other models and future events as part of the ongoing cycle of model improvement.

Incorporating prior work by reference in the paper is sufficient. Write what relevant information is contained in the prior work; then note its reference number in the summary table. Please verify that the whole chain of steps in referenced work is actually complete and up-to-date.

**This table and text has been adapted from Behrend et al. (2020).**

**Supplementary Text**

Literature searches were conducted for the period 01/11/2014 to 29/01/2019, according to the following outlined in Coral-Almeida et al. (2015):

In PubMed, using the Boolean operator AND, the terms “cysticercosis” AND “Taenia solium” AND “epidemiology” were introduced in the main search bar and the filters were activated for the period from 1988/12/31 to 2014/10/31.

In Web of Science, the strategy applied was introducing in the basic search bar the topic “cysticercosis” adding fields with the correspondent Boolean operators: AND Topic = (*Taenia solium*) AND Topic = (epidemiology).

In the Latin American & Caribbean Health Sciences Literature (LILACS) database, the strategy adopted consisted in introducing in the main search bar the terms “cysticercosis Taenia solium”. In the latter case the term “epidemiology” was excluded to obtain the maximum return of articles since LILACS is a smaller targeted database when compared to PubMed and Web Of Science.

In addition the literature databases searched in Coral-Almeida et al. (2015), the African Journals Online (AJOL) was also searched, with the search strategy terms including “cysticercosis *Taenia solium*”.

Following identification of article titles, the exclusion criteria at the title and abstract screening stage included: 1) wrong parasite species, 2) non-endemic countries, 3) only in humans, 4) pre-clinical/clinical research only, 5) diagnostic development only, 6) non-epidemiological study/ primary data not collected, and 7) unrelated topic.

At the full-text screening stage, the following inclusion criteria were applied: 1) community-based studies with a cross-sectional or longitudinal study design assessing *T. solium* human taeniasis (HTT) and human cysticercosis (HCC) prevalence; 2) community-based studies also assessing incidence rates; 3) diagnostics including copro-antigen and serology (immunoblot and ELISA) with sensitivity or specificity estimates available either from the broader literature or determined in the study; 4) intervention studies with baseline *T. solium* HTT and HCC prevalence data collected; 5) more than 2 age-categories with age-prevalence data presented; and 6) English, Spanish, Portuguese and French language inclusion. Where age was assessed as a risk factor or information on human age collected as described in the methods, attempts were made to contact the authors for access to the data. Equally, if age-(sero)prevalence profiles were incomplete, authors were contacted. A significant number of studies were excluded at full-text screening as the majority of studies were over 10 years old, and therefore either author contact information was out of date, not supplied, the authors did not reply when contacted, or authors could not find original data, and a number of full-texts were inaccessible after attempting to access through multiple sources (e.g. British Library).

__**Supplementary file references**__

Allan JC, Ávila G, Garcia Noval J, Flisser A, Craig PS. Immunodiagnosis of taeniasis by coproantigen detection. ***Parasitology*** 1990;**101** Pt 3:473–7. doi: 10.1017/s0031182000060686

Behrend MR, Basáñez MG, Hamley JID, Porco TC, Stolk WA, Walker M, de Vlas SJ; NTD Modelling Consortium. Modelling for policy: The five principles of the Neglected Tropical Diseases Modelling Consortium. ***PLoS Negl Trop Dis***. 2020 Apr 9;14(4):e0008033. doi: 10.1371/journal.pntd.0008033

Brandt JR, Geerts S, De Deken R, Kumar V, Ceulemans F, Brijs L, *et al*. A monoclonal antibody-based ELISA for the detection of circulating excretory-secretory antigens in *Taenia saginata* cysticercosis. ***Int J Parasitol*** 1992;**22**:471–7. doi: 10.1016/0020-7519(92)90148-e

Conlan J V, Vongxay K, Khamlome B, Dorny P, Sripa B, Elliot A, *et al*. A cross-sectional study of *Taenia solium* in a multiple taeniid-endemic region reveals competition may be protective***. Am J Trop Med Hyg*** 2012;**87**:281–91. doi: 10.4269/ajtmh.2012.11-0106.

Coral-Almeida M, Gabriel S, Abatih EN, Praet N, Benitez W, Dorny P. *Taenia solium* Human Cysticercosis: A Systematic Review of Sero-epidemiological Data from Endemic Zones around the World. ***PLoS Negl Trop Dis*** 2015;**9**:e0003919. doi: 10.1371/journal.pntd.0003919

DiagAutom. Human Cysticercosis IgG (T. Solium) ELISA Test kit. 2016. http://www.rapidtest.com/index.php?i=Parasitology-ELISA-kits&id=170&cat=17

Dorny P, Vercammen F, Brandt J, Vansteenkiste W, Berkvens D, Geerts S. Sero-epidemiological study of *Taenia saginata* cysticercosis in Belgian cattle. ***Vet Parasitol*** 2000;**88**:43–9. doi: 10.1016/s0304-4017(99)00196-x

Edia-asuke AU, Inabo HI, Mukaratirwa S, Umoh VJ, Whong CMZ, Asuke S, *et al*. Seroprevalence of human cysticercosis and its associated risk factors among humans in areas of Kaduna metropolis, Nigeria. ***J Infect Dev Ctries*** 2015;**9**:799–805. doi: 10.3855/jidc.5415

Fèvre EM, de Glanville WA, Thomas LF, Cook EAJ, Kariuki S, Wamae CN. An integrated study of human and animal infectious disease in the Lake Victoria crescent small-holder crop-livestock production system, Kenya. ***BMC Infect Dis*** 2017;17:457. doi: 10.1186/s12879-017-2559-6

Fleury A, Hernandez M, Avila M, Cardenas G, Bobes RJ, Huerta M, *et al*. Detection of HP10 antigen in serum for diagnosis and follow-up of subarachnoidal and intraventricular human neurocysticercosis. ***J Neurol Neurosurg Psychiatry*** 2007;**78**:970–4. doi: 10.1136/jnnp.2006.107243

Flórez Sánchez AC, Pastrán SM, Vargas NS, Beltrán M, Enriquez Y, Peña AP, *et al*. Cisticercosis en Colombia. Estudio de seroprevalencia 2008-2010. ***Acta Neurológica Colomb*** 2013;**29**(2):73–86. ISSN 0120-8748

García H, Gilman R, Gonzalez A, Verastegui M, Rodriguez S, Gadivia C, et al. Hyperendemic human and porcine *Taenia solium* infection in Perú. ***Am J Trop Med Hyg*** 2003;**68**:268–75. PMID: 12685628.

Gomes I, Veiga M, Embiruçu EK, Rabelo R, Mota B, Meza-Lucas A, *et al*. Taeniasis and cysticercosis prevalence in a small village from Northeastern Brazil. ***Arq Neuropsiquiatr*** 2002;**60**:93–103. doi: 10.1590/s0004-282x2002000200006

Harrison LJS, Joshua GWP, Wright SH, Parjhouse RME. Specific detection of circulating surface/secreted glycoproteins of viable cysticerci in *Taenia saginata* cysticercosis. ***Parasite Immunol*** 1989;**11**:351–70. doi:10.1111/j.1365-3024.1989.tb00673.x

Hancock K, Pattabhi S, Whitfield FW, Yushak ML, Lane WS, Garcia HH, *et al*. Characterization and cloning of T24, a *Taenia solium* antigen diagnostic for cysticercosis. ***Mol Biochem Parasitol*** 2006;**147**:109–17. doi: 10.1016/j.molbiopara.2006.02.004

Holt HR, Inthavong P, Khamlome B, Blaszak K, Keokamphe C, Somoulay V, *et al*. Endemicity of Zoonotic Diseases in Pigs and Humans in Lowland and Upland Lao PDR: Identification of Socio-cultural Risk Factors. ***PLoS Negl Trop Dis*** 2016;**12**:10(4):e0003913. doi: 10.1371/journal.pntd.0003913

Jayaraman T, Prabhakaran V, Babu P, Raghava MV, Rajshekhar V, Dorny P, *et al*. Relative seroprevalence of cysticercus antigens and antibodies and antibodies to *Taenia ova* in a population sample in south India suggests immunity against neurocysticercosis. ***Trans R Soc Trop Med Hyg*** 2011;**105**:153–9. doi: 10.1016/j.trstmh.2010.10.007

Kanobana K, Praet N, Kabwe C, Dorny P, Lukanu P, Madinga J, *et al*. High prevalence of *Taenia solium* cysticerosis in a village community of Bas-Congo, Democratic Republic of Congo. ***Int J Parasitol*** 2011;**41**:1015–8. doi: 10.1016/j.ijpara.2011.06.004

Lambert B. Chapter 8. A Student's Guide to Bayesian Statistics. London: SAGE Publications; 2018. p. 173–4. https://uk.sagepub.com/en-gb/eur/a-student's-guide-to-bayesian-statistics/book245409

Lescano AG, Garcia HH, Gilman RH, Gavidia CM, Tsang VCW, Rodriguez S, *et al*. *Taenia solium* Cysticercosis Hotspots Surrounding Tapeworm Carriers: Clustering on Human Seroprevalence but Not on Seizures. ***PLoS Negl Trop Dis*** 2009;**3**(1):e371. doi: 10.1371/journal.pntd.0000371

López M, Murillo C, Sarria N, Nicholls R, Corredor A. Estandarización y evaluación de la prueba de ELISA para la detección de anticuerpos en LCR y suero en neurocisticercosis. ***Acta Neurol Col*** 1988;**4**:164–9.

Madinga J, Kanobana K, Lukanu P, Abatih E, Baloji S, Linsuke S, *et al*. Geospatial and age-related patterns of *Taenia solium* taeniasis in the rural health zone of Kimpese , Democratic Republic of Congo. ***Acta Trop*** 2017;**165**:100–9. doi:10.1016/j.actatropica.2016.03.013

Moher D, Shamseer L, Clarke M, Ghersi D, Liberati A, Petticrew M, *et al*. Preferred Reporting Items for Systematic Review and Meta-Analysis Protocols (PRISMA-P) 2015 Statement. ***Syst Rev*** 2015;**1**:4(1):1. doi: 10.1186/2046-4053-4-1

Moro PL, Lopera L, Bonifacio N, Gilman RH, Silva B, Verastegui M, *et al*. *Taenia solium* infection in a rural community in the Peruvian Andes. ***Ann Trop Med Parasitol*** 2003;**97**:373–9. 10.1179/000349803235002371

Moyano LM, O'Neal SE, Ayvar V, Gonzalvez G, Gamboa R, Vilchez P, *et al*. High Prevalence of asymptomatic neurocysticercosis in an endemic rural community in Peru. ***PLoS Negl Trop Dis*** 2016;**10**:e0005130. doi: 10.1371/journal.pntd.0005130

Mwanjali G, Kihamia C, Kakoko DV, Lekule F, Ngowi H, Johansen MV, *et al*. Prevalence and risk factors associated with human *Taenia solium* infections in Mbozi District, Mbeya Region, Tanzania. ***PLoS Negl Trop Dis*** 2013;**7**:e2102. doi: 10.1371/journal.pntd.0002102

Mwape KE, Phiri IK, Praet N, Muma JB, Zulu G, van den Bossche P, *et al*. *Taenia solium* infections in a rural area of Eastern Zambia-A community based study. ***PLoS Negl Trop Dis*** 2012;**6**(3):e1594. doi: 10.1371/journal.pntd.0001594

Nguekam JP, Zoli AP, Zogo PO, Kamga ACT, Speybroeck N, Dorny P, *et al*. A seroepidemiological study of human cysticercosis in West Cameroon. ***Trop Med Int Health*** 2003;**8**:144–9. doi: 10.1046/j.1365-3156.2003.01000.x.

Noh J, Rodriguez S, Lee Y-M, Handali S, Gonzalez AE, Gilman RH, *et al*. Recombinant protein- and synthetic peptide-based immunoblot test for diagnosis of neurocysticercosis. ***J Clin Microbiol*** 2014;**52**:1429–34. doi: 10.1128/JCM.03260-13

Okello A, Ash A, Keokhamphet C, Hobbs E, Khamlome B, Dorny P, *et al*. Investigating a hyper-endemic focus of *Taenia solium* in northern Lao PDR. ***Parasit Vectors*** 2014;**28**:7:134. doi: 10.1186/1756-3305-7-134

Praet N, Rodriguez-Hidalgo R, Speybroeck N, Ahounou S, Benitez-Ortiz W, Berkvens D, *et al*. Infection with versus Exposure to *Taenia solium*: What Do Serological Test Results Tell Us? ***Am J Trop Med Hyg*** 2010;**83**:413–5. doi: 10.4269/ajtmh.2010.10-0121

Praet N, Verweij JJ, Mwape KE, Phiri IK, Muma JB, Zulu G, *et al*. Bayesian modelling to estimate the test characteristics of coprology, coproantigen ELISA and a novel real-time PCR for the diagnosis of taeniasis. ***Trop Med Int Heal*** 2013;**18**:608–14.

Sahlu I, Carabin H, Ganaba R, Preux PM, Cisse AK, Tarnagda Z, *et al*. Estimating the association between being seropositive for cysticercosis and the prevalence of epilepsy and severe chronic headaches in 60 villages of rural Burkina Faso. ***PLoS Negl Trop Dis*** 2019;**13**:e0007101.

Sato MO, Sato M, Yanagida T, Waikagul J, Pongvongsa T, Sako Y, *et al*. *Taenia solium*, *Taenia saginata*, *Taenia asiatica*, their hybrids and other helminthic infections occurring in a neglected tropical diseases' highly endemic area in Lao PDR. ***PLoS Negl Trop Dis*** 2018;**12**:e0006260. doi: 10.1371/journal.pntd.0006260

Theis JH, Goldsmith RS, Flisser A, Koss J, Chionino C, Plancarte A. Detection by immunoblot assay of antibodies to *Taenia solium* cysticerci in sera from residents of rural communities and from epileptic patients in Bali, Indonesia. ***Southeast Asian J Trop Med Public Health***. 1994;**25**(3):464-8. PMID: 7777908

Tsang VC, Brand JA, Boyer AE. An enzyme-linked immunoelectrotransfer blot assay and glycoprotein antigens for diagnosing human cysticercosis (*Taenia solium*). ***J Infect Dis*** 1989;**159**:50–9. doi: 10.1093/infdis/159.1.50

Wardrop NA, Thomas LF, Atkinson PM, de Glanville WA, Cook EAJ, Wamae CN, *et al*. The influence of socio-economic, behavioural and environmental factors on *Taenia* spp. transmission in Western Kenya: Evidence from a cross-sectional survey in humans and pigs. ***PLoS Negl Trop Dis*** 2015; **7:**9(12):e0004223. doi: 10.1371/journal.pntd.0004223.

Weka RP, Ikeh EI, Kamani J. Seroprevalence of antibodies (IgG) to *Taenia solium* among pig rearers and associated risk factors in Jos metropolis, Nigeria. ***J Infect Dev Ctries***. 2013;**15**:7(2):67-72. doi: 10.3855/jidc.2309

Wilkins PP, Allan JC, Verastegui M, Acosta M, Eason AG, Garcia HH, *et al*. Development of a serologic assay to detect *Taenia solium* taeniasis. ***Am J Trop Med Hyg*** 1999;**60**:199–204. doi: 10.4269/ajtmh.1999.60.199
